# Adding Different Exercise Training Volumes to a Diet-Induced Weight Loss Facilitates Fat Loss and Maintains Fat-Free Mass in a Dose-Depending Fashion in Persons with Newly Diagnosed Type 2 Diabetes: Secondary Findings from the DOSE-EX Multi-Arm, Parallel-Group, Randomized Trial

**DOI:** 10.1101/2023.06.04.23290749

**Authors:** Mark P.P. Lyngbæk, Grit E. Legaard, Nina S. Nielsen, Cody Durrer, Thomas P. Almdal, Morten Asp Vonsild Lund, Benedikte Liebetrau, Caroline Ewertsen, Carsten Lauridsen, Thomas P.J. Solomon, Kristian Karstoft, Bente K. Pedersen, Mathias Ried-Larsen

## Abstract

**OBJECTIVE:** To assess the dose-response effects of exercise in combination with a diet-induced weight loss on fat mass (FM) percentage (FM%) in persons with diagnosed type 2 diabetes.

**RESEARCH DESIGN AND METHODS:** In this secondary analysis of a four-armed randomized trial (Clinicaltrials.gov NCT03769883) 82 persons (35% females, mean age and standard deviation (SD) 58.2 (9.8) years) living with type 2 diabetes were randomly allocated to the control group (N=21, CON), diet control (25% energy restriction; *N*=20, DCON), diet control and exercise three times/week (two sessions of aerobic and one session combining resistance and aerobic training; *N*=20, MED), or diet control and exercise six times/week (four sessions of aerobic and two sessions combining resistance and aerobic training; *N*=21, HED) for 16 weeks. The primary outcome was the change in FM percentage points (pp). Secondary outcomes included fat-free mass and visceral adipose tissue volume.

**RESULTS:** Type 2 diabetes duration was 4.0 years (interquartile range 1.9 to 5.5), body weight (SD) 101.4 kg (14.6), FM% (SD) 39.4 (6.7). FMpp decreased compared to standard care −3.5 pp (95% CI −5.6 to −1.4) p=0.002, −6.3 pp (CI −8.4 to −4.1) p<0.001, and −8.0 pp (95% CI −10.2 to −5.8) p<0.001, for DCON, MED, and HED, respectively. The difference between HED and MED was −1.8 pp [95% CI −3.9 to 0.4]; p=0.11).

**CONCLUSIONS:** All interventions were superior in reducing FMpp compared to standard care in a dose-dependent manner. Adding three or six sessions of exercise to a low-calorie diet was superior in reducing FM compared to a low-calorie diet alone.

**Article Highlights:**

a. Why did we undertake this study? Exercise and weight loss are recommended for persons with type 2 diabetes. It is unclear if adding exercise, and which amount of exercise, to a low-calorie diet supports additional fat mass loss.
b. What is the specific question(s) we wanted to answer? What is the dose-response effect of exercise combined with a moderate caloric restriction on changes in fat mass?
c. What did we find? Adding exercise to a diet-induced weight loss reduced fat mass and preserved fat-free mass in a dose-dependent manner.
d. What are the implications of our findings? Adding exercise to a moderate caloric restriction dose-dependently facilitates reductions in fat mass by enlarging weight loss and fat loss.

## Introduction

Excessive adiposity or overweight/obesity are risk factors for the onset of type 2 diabetes [1, 2] and approximately 90% of persons with type 2 diabetes have overweight or obesity [3]. Hence, the current recommendations for the management of hyperglycemia in type 2 diabetes define weight loss as a treatment target. Importantly, it is the net negative energy balance enabling the reduction in excessive amounts of fat mass (FM) that is the major therapeutic aim of weight loss [2, 4–9].

Despite the benefits of weight loss, there is a concomitant risk of losing up to 50% fat-free mass (FFM) or lean body mass from the total weight loss [6, 7, 10]. Besides not losing fat during weight loss, a long-term consequence of losing FFM may be an increased risk of sarcopenia and/or physical impairment already increased in people living with type 2 diabetes [8, 11]. It has been proposed that a bidirectional relationship between type 2 diabetes severity and duration, and sarcopenia exists [11]. Consequently, interventions that facilitate a reduction in excessive adiposity while preserving FFM are clinically warranted.

Exercise may preserve FFM during calorie restriction [7, 12, 13], and even facilitate reductions in FM beyond what is expected from calorie restriction alone [14–16], but others suggest that exercise combined with calorie restriction does not decrease FM more than calorie restriction alone when the weight loss approaches ≥5-10% body weight [17–19] and therefore question the utility of exercise in reducing excessive adiposity.

The current physical activity recommendations for persons living with type 2 diabetes do not include considerations for optimizing fat loss while preserving FFM *during* weight loss [8, 20]. Moreover, the optimal and/or sufficient exercise dose to elicit such changes is unknown. Thus, we performed a secondary analysis of the Dose-Ex study [21] with the primary aim of testing the hypothesis that FM% (expressed as changes in FM percentage points [FMpp]) decreases in an exercise dose-dependent manner when combined with caloric restriction compared to caloric restriction and/or standard care only during 16 weeks. Secondarily, we aimed to investigate the effect of the intervention on FFM and visceral adipose tissue (VAT).

### Research Design and Methods Study design and Participants

This study is a secondary analysis of a randomized clinical trial [21] conducted between February 2019 and October 2021 at the Centre for Physical Activity Research (CFAS), Rigshospitalet, Copenhagen, Denmark. The study was pre-registered at www.clinicaltrials.gov (NCT03769883) and approved by the Scientific Ethical Committee of the Capital Region of Denmark (approval number H-18038298). All participants provided oral and written informed consent before any testing. Guidelines from the Helsinki Declaration were followed. Data on this secondary analysis are reported following the CONSORT for multi-arm trials (supplementary table S1). A full description of the study protocol has been published elsewhere [22]. A statistical analysis plan was published on the 24^th^ ^of^ November 2022 before commencing any of the inherent analyses [23].

The main inclusion criteria were Caucasian male and female aged 18–80 years, diagnosed with type 2 diabetes <7 years, physically inactive, no weight loss of >5 kg within the past 6 months, BMI >27 kg/m^2^, and <40 kg/m^2^.

### Interventions

The 16-week intervention included three components: standard care, exercise, and diet-induced weight loss. *The Control Group (CON)* received standard care and was encouraged to maintain their habitual physical activity and dietary habits throughout the study. *The Diet Control Group (DCON)* received standard care and a diet intervention aiming at a weight loss through ∼25-30% energy deficit/day based on the age-adjusted Oxford equation [24] with a macro-nutrient distribution of 45–60E% carbohydrate, 15–20E% protein, and 20–35E% fat (<7E% saturated fat). *The Moderate Exercise Dose Group (MED)* received standard care, diet intervention, and an exercise intervention with two aerobic training sessions and one combined aerobic and resistance training session/per week. In total 150-165 minutes of weekly exercise. *The High Exercise Dose Group (HED)* received the standard care and diet interventions with a total of four aerobic training sessions/per week and two sessions/per week with combined aerobic training and resistance training. In total 300-330 minutes of weekly exercise (i.e., twice the exercise volume as MED). Maximal heart rate (HRmax) was determined at baseline VO2max testing. Target-intensity of 60–100% was HRmax for aerobic training and resistance training was performed as 8-12 repetitions with proximity to failure of 0-3 repetitions-in-reserve on the large muscle groups. All participants were encouraged to consume a 100-200 kcal snack before and after each exercise session to avoid feelings of hypoglycemia [22]. All heart rate profiles (Polar V800, Polar, Holte, DK), repetitions-in-reserve sets, and modifications of exercise selections were recorded during the exercise interventions and all training sessions were supervised by educated trainers. Data on the diet - and exercise intervention have been reported previously [21].

### Outcomes

*The primary outcome* was the change in FMpp from baseline to the 16-week follow-up assessed with Dual-energy X-ray absorptiometry (DXA).

*The secondary outcomes* were changes in FM kg, FFM kg, and FFMpp assessed with DXA, and VAT cm^3^ assessed with Magnetic Resonance Imaging (MRI) from baseline to the 16-week follow-up. Weight loss was measured with an electronic scale and height was measured with a Holtain stadiometer (Holtain, Crymych, UK) according to standard procedures.

*The post hoc outcomes* were changes in appendicular FFM kg, appendicular FFMpp, FM:FFM ratio assessed with DXA, abdominal subcutaneous adipose tissue cm^3^ (aSAT), VAT:aSAT ratio from baseline to the 16-week follow-up. Body weight, beta-cell disposition index, insulin sensitivity index, and insulin secretion have been reported previously [21].

### Body composition assessments

A DXA and an MRI scan were conducted at least one week apart but within a two-week window at baseline and at 16-week follow-up. The DXA was a Prodigy Advance, GE Medical Systems Lunar, Madison, WI, USA. The participants were instructed to maintain their diet and refrain from exercise 48 hours before testing. The participants arrived at the testing facility at 07.30 AM after an overnight fast (≥10 hours) and were asked to void before the DXA scan. All DXA scans were manually adjusted, when appropriate, and reviewed in agreement with the DXA Encore Software bone densitometer parameters by one investigator. The participants had fasted for 5 hours before the MRI.

Volumes for VAT and aSAT were calculated from MR images (Siemens Magnetom Prisma 3 Tesla matrix magnetic resonance scanner Erlangen, Germany). VAT and aSAT were assessed from transverse slices between the intervertebral discs separating S1/L5 and Th12/L1. T1-weighted images were acquired during an end-expiration breath-hold (TR = 5.43 ms, TE = 1.23 ms, flip angle = 9°), with a slice thickness of 3 mm. Based on the in-phase and opposed-phase images, the Slice-O-Matic software [25] was used to generate fat and water-only images; these images were then analyzed for VAT and aSAT surface area, multiplied by the slice thickness, and summed to calculate total volume. All scans were analyzed by one investigator. The total number of MRI and DXA scans and deviations from the analyses are reported (Supplementary Table S9).

### Insulin sensitivity, insulin secretion, and beta-cell function

A 3-stage hyperglycemic clamp and 3-hour mixed meal tolerance test were performed at baseline and at 16-week follow-up (detailed procedures published in [22]). The results for insulin sensitivity, insulin secretion rate, and beta-cell disposition index have been reported previously [21] and are used in this study to investigate the mediation of changes in body composition on glucose homeostasis.

### Randomization and blinding

A computer-generated randomization schedule in a ratio of 1:1:1:1, using permuted block sizes and stratified by sex, was prepared by an independent person not located at the testing facility. After baseline measurements, the study nurse informed the participant of group allocation. Personnel involved in data collection were blinded to participant allocation.

### Sample size and power considerations

The sample size was based on the late-phase DI [22]. A total sample size of 80 participants in the per-protocol population (approximately 20 participants in each group) would yield a statistical power of 87.7%.

### Statistical Methods

The statistical analyses were performed using R [26]. Continuous data, including the primary, secondary, and exploratory outcomes, were analyzed using constrained baseline longitudinal analyses via linear mixed models [27]. The model included the fixed effects for time (2 levels), treatment (coded 0 for all groups at baseline and coded 0, 1, 2, or 3 at follow-up for the CON, DCON, MED, and HED, respectively), sex (2 levels) and the unique patient identifier as a random intercept. The lme4 package (v1.1-3) was used to construct the linear mixed-models for the analysis [28]. The potentially biased *per-protocol* population analysis was further adjusted for putative confounders: diabetes duration and baseline maximal oxygen consumption (ml O2/kg/min). Data are presented as post-intervention estimated marginal means (for each treatment allocation) and the difference in the mean changes from baseline with 95% confidence intervals (95% CI) unless stated otherwise. Estimated marginal means and contrasts were calculated via the emmeans package [29]. If the global test indicated between-group differences in the treatment effect (H0,DCON = H0,MED = H0,HED = H0,CON; p≤0.1), pairwise between-group differences, in the order described above, were explored. The model assumptions were assessed visually via normal probability plots and residuals vs. fitted values plots. Variables not meeting the model assumptions were log-transformed using the natural logarithm and estimated marginal means were back-transformed to their original scale; in this case, mean differences are presented as the ratio of geometric means and interpreted as the relative change from baseline to follow-up between the groups (performed for the variables FFM kg and FFM/FM ratio). If a log-transformation of the data did not satisfy model assumptions, non-parametric bootstrap analysis using 2000 resamples with replacement was performed, and bias-corrected and accelerated 95% confidence intervals were calculated (performed for the variables VAT, aSAT, and VAT:aSAT ratio). By default, no imputations were used (statistical or otherwise) for the analysis, and any missing data were assumed to be missing at random.

Linear trends (interpreted as a linear dose-response relationship) were examined by converting the treatment category to a continuous variable in the main model (i.e., 0, 1, 2, and 3 for CON, DCON, MED, and HED, respectively) and tested using a type II Wald-test (p-value reported). Linearity was inspected visually, and the p-for-trend was only calculated for the primary and secondary outcomes if the treatment and outcome displayed a linear relationship.

If there was a significant treatment effect on an assessment of body composition, a statistical mediation analysis was performed to examine the extent to which the observed treatment effect (in the intervention groups) on the primary and secondary outcomes from the main paper (late-phase disposition index, late-phase insulin sensitivity, late-phase insulin secretion, the oral disposition, and insulin sensitivity indices) was mediated by the change in that body composition compartment (body fat percentage, fat-free mass percentage, subcutaneous and visceral adipose tissue percentage). This simple mediation analysis partitions the total causal effect into average direct effects (ADE) and average causally mediated effects (ACME; otherwise known as indirect effects). Bias-corrected and accelerated 95% confidence intervals were generated via non-parametric bootstrap analysis (5000 resamples with replacement). The statistical significance level (for superiority) was set at α<0.05 (two-sided).

## Data and Resource Availability

Data are not available for download due to privacy/ethical restrictions under the EU GDPR. Specific requests for access to the trial and unique biological data as well as code may be sent to the corresponding author. Based on the request access may be provided to a named individual in agreement with the rules and regulations of the Danish Data Protection Agency and the National Committee on Health Research Ethics.

## Results

### Participant characteristics and adherence

Eighty-two persons were included (Fig. 1) and five (six percent) participants were lost-to-follow-up. The mean (standard deviation [SD]) age was 58.2 years (9.8), BMI was 33 kg/m^2^ (3.7), and glycated hemoglobin was 50.2 mmol/mol (6.6). Twenty-nine percent were females, and the median and interquartile range (IQR) type 2 diabetes duration was 4.0 years (1.9 to 5.5). Baseline characteristics are presented in Table 1. Mean (SD) adherence to the dietary intervention was 92% (11) for DCON, 91% (18) for MED, and 88% (13) for HED. The mean (SD) reduction in total daily energy intake (kilocalories/day) was DCON −708 (452), MED - 509 (424), HED - 620 (528), and CON - 207 (495) and the macronutrient distribution was overall similar between intervention groups (Supplementary table S2). The mean (SD) exercise adherence was 93% (18) and 86% (28) and for MED and HED, respectively (Supplementary tables S3-8).

**Figure 1.**
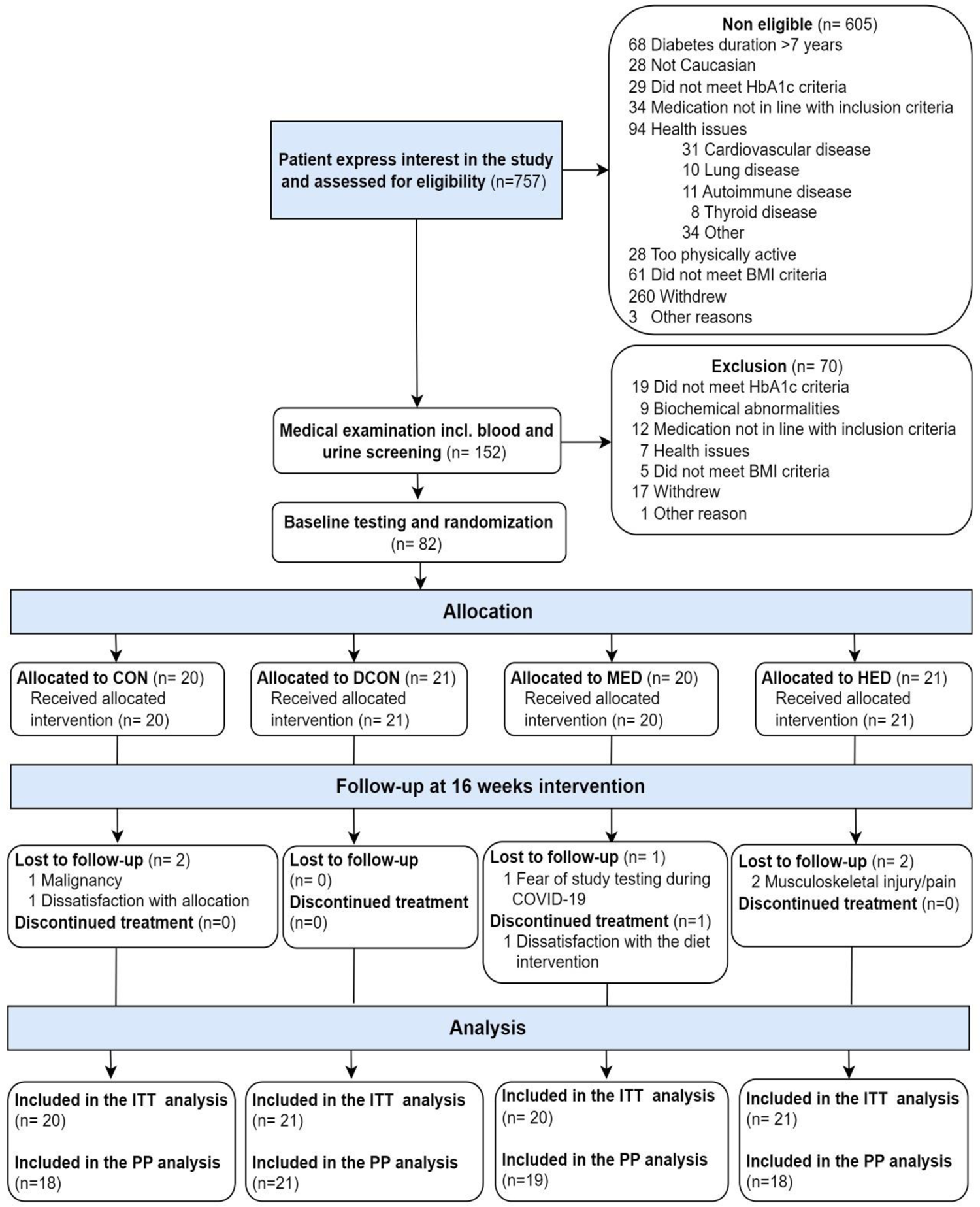
Flow of participants. CON: Control group, DCON: Diet control group, MED: Moderate exercise dose, HED: High exercise dose, ITT: intention-to-treat, PP: per-protocol.

**Table 1.** Baseline characteristics. . Data are presented as mean (SD) or median (IQR, meaning p25 to p75). CON, control group, DCON: Diet control group: MED: Moderate volume exercise, HED: High volume exercise, HbA1c: glycated hemoglobin A1c, LDL-C: low-density lipoprotein cholesterol, BMI: body mass index (calculated as weight in kilograms divided by height in meters squared). SLGT2i: selective sodium glucose co-transporter 2 inhibitors, DPP4i: dipeptidyl peptidase 4 inhibitors, ARB: angiotensin II receptor blockers, ACEi: angiotensin-converting enzyme inhibitors, CCB: calcium channel blockers. FFM/FM: fat-free mass/ fat mass, VAT/aSAT: visceral adipose tissue/ abdominal subcutaneous adipose tissue. Energy intake was assessed via a self-reported 3-day dietary diary and blood pressure was assessed by 3-day home blood pressure monitoring as described in the primary paper. ^#^n=20; ^##^n=19, ^###^n=18.

|  | CON | DCON | MED | HED | Total |
| --- | --- | --- | --- | --- | --- |
|  | n=20 | n=21 | n=20 | n=21 | n=82 |
|  | Mean (SD) | Mean (SD) | Mean (SD) | Mean (SD) | Mean (SD) |
| **Age (years)** | 59.1 (9.2) | 55.9 (10.0) | 60.9 (7.6) | 57.3 (11.8) | 58.2 (9.8) |
| **Sex (N (%) female)** | 7.0 (35.0) | 7.0 (33.3) | 7.0 (35.0) | 8.0 (38.1) | 29.0 (35.4) |
| **Type 2 diabetes duration (years)** | 3.5 (1.2) | 3.9 (2.9) | 4.0 (2.1) | 4.2 (3.4) | 4.0 (1.9) |
| **Glycemic control** |  |  |  |  |  |
| HbA1c (mmol/mol) | 52 (7) | 49 (7) | 51 (6) | 49 (7) | 50 (7) |
| HbA1c (%) | 6.9 (0.6) | 6.7 (0.7) | 6.8 (0.6) | 6.7 (0.6) | 6.7 (0.6) |
| Fasting glucose, median (IQR), (mmol/l) | 9.1 (7.3 ; 10.3) | 7.8 (7.3 ; 9.8) | 7.7 (7.1 ; 9.6) | 7.8 (7.0 ; 9.6) | 7.8 (7.1 ; 10.0) |
| **Blood lipids** |  |  |  |  |  |
| LDL-C (mmol/L) | 3.2 (0.8) | 2.9 (0.7) | 2.9 (0.6) | 2.6 (0.6) | 2.9 (0.7) |
| Fasting triglycerides (mmol/L) | 1.5 (1.3) | 1.5 (1.2) | 1.8 (1.3) | 1.4 (1.1) | 1.5 (1.1) |
| **Blood pressure** |  |  |  |  |  |
| Systolic (mmHg) | 128 (12) | 127 (10.3<sup>#</sup>) | 133 (11.2<sup>##</sup>) | 129 (11) | 129 (11) |
| Diastolic (mmHg) | 78 (6) | 80 (6.7<sup>#</sup>) | 81 (7.5<sup>##</sup>) | 78 (7) | 79 (7) |
| **Glucose-lowering medication, N (%)** |  |  |  |  |  |
| None | 5.0 (25.0) | 8.0 (38.1) | 5.0 (25.0) | 5.0 (23.8) | 23.0 (28.0) |
| Biguanide | 9.0 (45.0) | 7.0 (33.3) | 11.0 (55.0) | 9.0 (42.9) | 36.0 (43.9) |
| Biguanide + SGLT2i or DPP4i | 5.0 (25.0) | 5.0 (23.8) | 4.0 (20.0) | 6.0 (28.6) | 20.0 (24.4) |
| Biguanide + SGLT2i + DPP4i | 1.0 (5.0) | 1.0 (4.8) | 0.0 (0.0) | 1.0 (4.8) | 3.0 (3.7) |
| **Lipid-lowering medication, No (%)** |  |  |  |  |  |
| None | 7.0 (35.0) | 7.0 (33.3) | 9.0 (45.0) | 5.0 (23.8) | 28.0 (34.1) |
| Statin | 13.0 (65.0) | 14.0 (66.7) | 11.0 (55.0) | 16.0 (76.2) | 54.0 (65.9) |
| **Blood pressure-lowering medication, No (%)** |  |  |  |  |  |
| None | 11.0 (55.0) | 9.0 (42.9) | 11.0 (55.0) | 6.0 (28.6) | 37.0 (45.1) |
| ARB or ACEi | 4.0 (20.0) | 5.0 (23.8) | 4.0 (20.0) | 6.0 (28.6) | 19.0 (23.2) |
| ARB or ACEi + Thiazide or CCB | 4.0 (20.0) | 4.0 (19.0) | 4.0 (20.0) | 6.0 (28.6) | 18.0 (22.0) |
| ARB or ACEi + Thiazide + CCB | 1.0 (5.0) | 3.0 (14.3) | 1.0 (5.0) | 3.0 (14.3) | 8.0 (9.8) |
| **Physical fitness** |  |  |  |  |  |
| Absolute VO<sub>2</sub> max (ml/min) | 2445.6 (413.2) | 2611.6 (618.2) | 2512.9 (634.5) | 2582.0 (714.4) | 2539.5 (599.2) |
| Relative VO<sub>2</sub> max (ml/kg/min) | 24.5 (3.7) | 25.7 (3.6) | 24.7 (4.6) | 24.8 (4.3) | 24.9 (4.0) |
| Watt max | 192.8 (41.3) | 204.8 (48.5) | 189.7 (49.6) | 204.8 (66.6) | 198.2 (52.0) |
| 1 RM chest press, median (IQR), (kg) | 40.0 (35.0 ; 55.0<sup>##</sup>) | 47.5 (35.0 ; 57.5) | 45.0 (35.0 ; 57.5<sup>###</sup>) | 52.5 (25.0 ; 65.0) | 45.0 (35.0 ; 57.5) |
| 1 RM leg extension (kg) | 68.0 (20.9<sup>#</sup>) | 75.2 (24.4<sup>#</sup>) | 63.6 (17.3) | 68.0 (22.4) | 68.7 (21.4) |
| **Body composition (absolute values)** |  |  |  |  |  |
| Body weight (kg) | 100.3 (12.3) | 100.8 (15.4) | 101.6 (16.1) | 102.8 (15.2) | 101.4 (14.6) |
| BMI (kg/m<sup>2</sup>) | 32.4 (3.6) | 33.2 (3.8) | 33.2 (4.1) | 33.4 (3.5) | 33.1 (3.7) |
| Fat mass (kg) | 38.0 (8.3) | 37.2 (7.9) | 38.3 (10.2) | 39.6 (7.9) | 38.3 (8.5) |
| Fat-free mass (kg) | 61.3 (10.4) | 62.9 (12.4) | 62.4 (11.1) | 62.7 (11.5) | 62.3 (11.2) |
| Leg fat-free mass (kg) | 20.4 (3.6) | 20.7 (4.5) | 20.1 (4.0) | 20.6 (4.3) | 20.5 (4.1) |
| Visceral adipose tissue (cm<sup>3</sup>) | 3870.1<br>(1273.4) | 4659.5<br>(1719.8) | 4937.0<br>(1220.4) | 4257.9<br>(1006.8) | 4451.3 (1368.0) |
| Abdominal subcutaneous adipose tissue (cm<sup>3</sup>) | 5206.6<br>(1828.6) | 4660.2<br>(686.3) | 3414.1<br>(504.2) | 4127.6<br>(1341.4) | 4378.0 (1359.8) |
| **Body composition (relative to body weight)** |  |  |  |  |  |
| Fat mass (%) | 39.7 (7.2) | 38.7 (6.7) | 39.1 (7.2) | 40.1 (6.2) | 39.4 (6.7) |
| Fat-free mass (%) | 63.8 (7.4) | 64.7 (6.8) | 64.2 (7.5) | 63.2 (6.5) | 64.0 (6.9) |
| Leg fat-free mass (%) | 33.3 (1.4) | 32.9 (1.4) | 32.1 (2.3) | 32.8 (1.9) | 32.8 (1.8) |
| FFM/FM ratio | 1.7 (1.4-2.0) | 1.6 (1.4-2.1) | 1.8 (1.3-2.1) | 1.6 (1.3-1.8) | 1.7 (1.4-2.1) |
| VAT/aSAT ratio | 0.8 (0.4) | 0.7 (0.3) | 1.5 (0.3) | 1.1 (0.4) | 1.0 (0.4) |
| **Diet** |  |  |  |  |  |
| Energy intake, median (IQR), (kcal/day) | 1976.5<br>(1772.0 ; 2485.0) | 1990.0<br>(1843.0 ; 2247.0) | 2052.0<br>(1800.0 ; 2389.0<sup>##</sup>) | 2052.0<br>(1571.0 ; 2586.0) | 1995.0 (1784.0 ; 2465.0) |
| **Free-living physical activity level** |  |  |  |  | (N=81) |
| Valid days (N) | 6.0 (1.4) | 6.4 (1.3) | 5.8 (1.1) | 6.0 (1.6) | 6 (6 ; 7) |
| Non-wear time (min/day) | 0 (0 ; 5) | 0 (0 ; 0) | 0 (0 ; 0) | 0 (0 ; 0) | 0 (0 ; 0) |
| Total physical activity (counts per minute) | 291 (215 ; 376) | 276 (224 ; 366) | 276 (168 ; 369) | 303 (226 ; 340) | 290 (217 ; 366) |
| Moderate and vigorous physical activity (minutes/day) | 17 (11 ; 24) | 15 (6 ; 20) | 15 (4 ; 27) | 14 (8 ; 25) | 15 (6 ; 25) |
| Sitting time (minutes/day) | 529 (484 ; 611) | 592 (515 ; 654) | 635 (429 ; 684) | 580 (514 ; 671) | 569 (483 ; 666) |
| Stepping (steps/day) | 5499 (3561 ; 7452) | 4303 (3855 ; 5798) | 4034 (2953 ; 6216) | 4380 (3336 ; 7110) | 4493 (3336 ; 6570) |

### Primary outcome

#### Fat mass percentage

All intervention groups decreased in FMpp compared to CON in a dose-depended manner (linear trend p<0.0001) (HED vs. CON −8.0pp [95% CI −10.2 to −5.8]; p<0.001, MED vs. CON −6.3pp [95% CI −8.4 to −4.1]; p<0.001, DCON vs. CON −3.5pp [95% CI −5.6 to −1.4]; p=0.002). MED and HED decreased more compared to DCON (MED vs. DCON −2.8pp [95% CI −4.9 to −0.7]; p=0.01, and HED vs. DCON −4.5pp [95% CI −6.6 to −2.4]; p<0.001). The difference in FMpp between HED and MED was −1.8pp [95% CI −3.9 to 0.4]; p=0.11) (Table 2 and Fig. 2).

**Figure 2.**
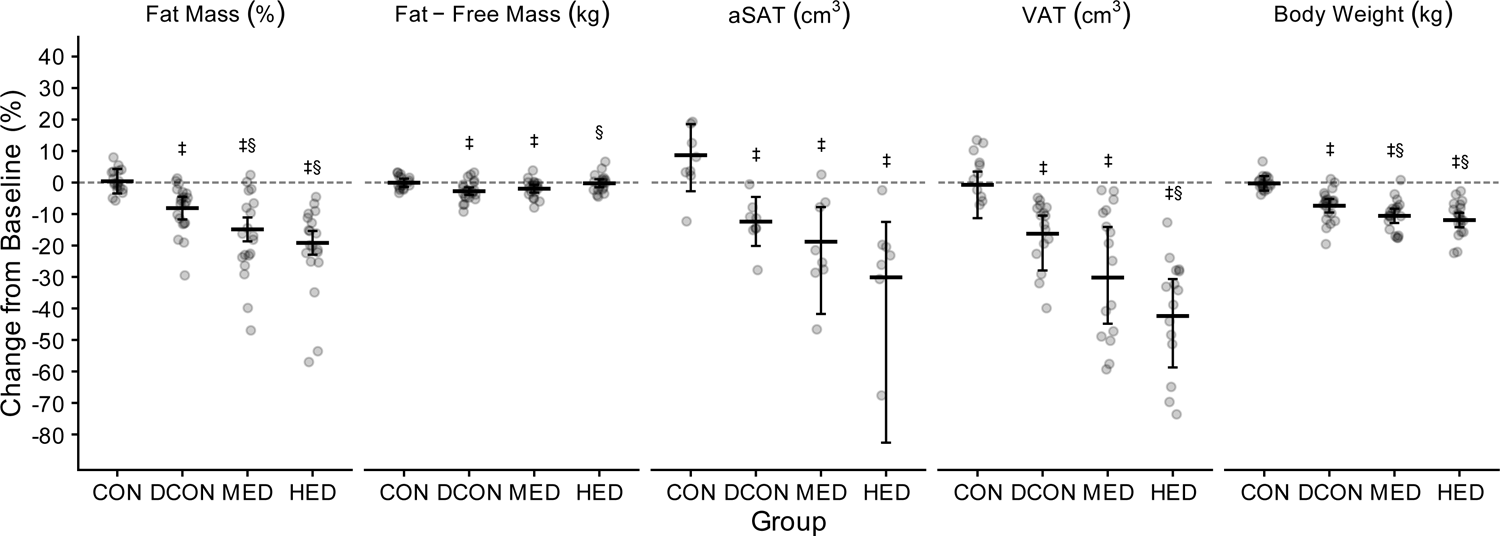
Changes from constrained baseline to 16 weeks follow-up in the primary and secondary outcomes. Data are presented as marginal means (bar charts overlaid with individual values) with 95% confidence intervals. Data were analyzed using a constrained baseline longitudinal model via linear mixed models. Results are adjusted for sex. *Mean difference expressed as a percent difference (ratio of geometric means) from the log-transformed analysis. †Bias-corrected and accelerated confidence intervals derived from non-parametric bootstrap analysis (in this case a p-value can’t be obtained). ‡Statistically significant difference compared to CON; §Statistically significant difference compared to DCON; Statistically significant differences are defined as p<0.05 or a bias-corrected and accelerated confidence interval that does not include 0. FFM (kg)*: fat-free mass, FM (%): fat mass, aSAT (cm^3^)†: abdominal subcutaneous adipose tissue, VAT (cm^3^)†: Visceral adipose tissue, Body weight (kg), CON: control group, DCON: Diet control group, MED: Moderate exercise dose, HED: High exercise dose.

**Table 2.** Pairwise comparisons of the change in the primary outcome and secondary outcomes. Data are adjusted mean differences and 95% confidence intervals derived from constrained baseline longitudinal analysis via linear mixed models. *Mean difference expressed as a percent difference (ratio of geometric means) from the log-transformed analysis. †Bias-corrected and accelerated confidence intervals derived from non-parametric bootstrap analysis (in this case a p-value can’t be obtained). MD: Mean difference, CI: confidence intervals, FFM: fat-free mass, FM: fat mass, aSAT: abdominal subcutaneous adipose tissue, VAT: Visceral adipose tissue, CON: control group, DCON: Diet control group, MED: Moderate exercise dose, HED: High exercise dose.

|  | HED vs. CON |  | MED vs. CON |  | DCON vs. CON |  | HED vs. DCON |  | MED vs. DCON |  | HED vs. MED |  | Global P |
| --- | --- | --- | --- | --- | --- | --- | --- | --- | --- | --- | --- | --- | --- |
|  | MD (95% CI) | P | MD (95% CI) | P | MD (95% CI) | P | MD (95% CI) | P | MD (95% CI) | P | MD (95% CI) | P |  |
| **Primary outcome** |  |  |  |  |  |  |  |  |  |  |  |  |  |
| Fat mass (pp) | -8.0 (-10.2 to -5.8) | <0.001 | -6.3 (-8.4 to -4.1) | <0.001 | -3.5 (-5.6 to -1.4) | 0.002 | -4.5 (-6.6 to -2.4) | <0.001 | -2.8 (-4.9 to -0.7) | 0.01 | -1.8 (-3.9 to 0.4) | 0.11 | <0.001 |
| **Secondary outcomes** |  |  |  |  |  |  |  |  |  |  |  |  |  |
| Body weight (kg) | -11.6 (-14.8 to -8.4) | <0.001 | -10.3 (-13.5 to -7.1) | <0.001 | -7.1 (-10.2 to -3.9) | <0.001 | -4.5 (-7.6 to -1.4) | 0.005 | -3.2 (-6.3 to -0.1) | 0.04 | -1.3 (-4.5 to 1.8) | 0.41 | <0.001 |
| Body weight (% difference) | -11.7 (-14.9 to -8.4) | <0.001 | -10.3 (-13.6 to -7.1) | <0.001 | -7.1 (-10.3 to -4.0) | <0.001 | -4.9 (-8.3 to -1.5) | 0.005 | -3.5 (-6.8 to -0.1) | 0.04 | -1.5 (-5.1 to 2.1) | 0.41 | <0.001 |
| Fat mass (kg) | -11.4 (-14.1 to -8.6) | <0.001 | -9.0 (-11.8 to -6.3) | <0.001 | -5.6 (-8.3 to -2.9) | <0.001 | -5.8 (-8.4 to -3.1) | <0.001 | -3.4 (-6.1 to -0.8) | 0.01 | -2.3 (-5.0 to 0.3) | 0.09 | <0.001 |
| Fat-free mass (kg) (% difference) | -0.2 (-2.0 to 1.7)* | 0.86 | -1.9 (-3.7 to -0.1)* | 0.04 | -2.7 (-4.4 to -0.9)* | 0.003 | 2.6 (0.8 to 4.4)* | 0.005 | 0.8 (-0.9 to 2.6)* | 0.36 | 1.8 (-0.0 to 3.6)* | 0.06 | 0.004 |
| Fat-free mass (pp) | 8.4 (6.1 to 10.6) | <0.001 | 6.6 (4.4 to 8.9) | <0.001 | 3.7 (1.5 to 5.9) | 0.001 | 4.7 (2.5 to 6.8) | <0.001 | 2.9 (0.7 to 5.1) | 0.010 | 1.8 (-0.5 to 4.0) | 0.12 | <0.001 |
| VAT (cm<sup>3</sup>) | -1786.4 (-2380.5 to -1131.5)† | NA | -1264.0 (-2129.3 to -725.2)† | NA | -666.0 (-1109.4 to -313.1)† | NA | -1120.4 (-1664.9 to -332.2)† | NA | -598.1 (-1444.1 to -9.0)† | NA | -522.4 (-1363.6 to 402.7)† | NA | NA |
| **Post hoc outcomes** |  |  |  |  |  |  |  |  |  |  |  |  |  |
| aSAT (cm<sup>3</sup>) | -1714.8 (-3896.0 to -693.4)† | NA | -1215.1 (-2182.1 to -539.5)† | NA | -933.5 (-1604.9 to -483.2)† | NA | -781.3 (-3114.5 to 64.6)† | NA | -281.7 (-1316.0 to 346.4)† | NA | -499.6 (-3061.5 to 392.1)† | NA | NA |
| Leg fat-free mass (kg) (% difference) | 1.5 (-1.9 to 5.0) | 0.39 | 1.4 (-1.9 to 4.9) | 0.40 | -2.3 (-5.5 to 0.9) | 0.16 | 3.9 (0.6 to 7.3) | 0.02 | 3.9 (0.5 to 7.3) | 0.02 | 0.0 (-3.2 to 3.4) | 0.98 | 0.06 |
| Leg fat-free mass (pp) | 0.4 (-0.5 to 1.3) | 0.43 | 0.7 (-0.2 to 1.6) | 0.15 | -0.0 (-0.9 to 0.8) | 0.93 | 0.4 (-0.5 to 1.3) | 0.36 | 0.7 (-0.2 to 1.6) | 0.11 | -0.3 (-1.2 to 0.6) | 0.51 | 0.34 |
| FFM/FM | 45.9 (30.3 to 63.5)* | <0.001 | 34.9 (20.4 to 51.1)* | <0.001 | 17.4 (5.1 to 31.2)* | 0.005 | 24.2 (11.4 to 38.6)* | <0.001 | 14.8 (2.9 to 28.1)* | 0.01 | 8.2 (-3.3 to 21.0)* | 0.17 | <0.001 |
| VAT/aSAT (% difference) | -6.0 (-33.4 to 13.5)† | NA | -25.3 (-73.3 to 18.5)† | NA | 1.7 (-10.3 to 18.4)† | NA | -7.6 (-27.2 to 18.7)† | NA | -26.6 (-82.8 to 6.3)† | NA | 25.9 (-30.9 to 96.3)† | NA | NA |

### Secondary outcomes

#### Body weight change

All intervention groups decreased in weight compared to CON (p<0.001 for all comparisons) in a dose-dependent manner (linear trend p<0.0001). MED and HED decreased compared to DCON. There was no difference in weight loss between HED and MED (Table 2 and Fig 2.)

#### Fat mass change

All intervention groups decreased FM kg compared to CON in a dose-dependent manner (linear trend p<0.0001) by −5.6 kg [95% CI −8.3 to −2.9], −9.0 kg [95% CI − 11.8 to −6.3], −11.4 kg [95% CI −14.1 to −8.6] for DCON, MED, and HED, respectively (p<0.001 for all comparisons). MED and HED decreased FM kg more compared to DCON (MED vs. DCON −3.4 kg [95% CI −6.1 to −0.8]; p=0.01, and HED vs. DCON −5.8 kg [95% CI −8.4 to − 3.1]; p<0.001). The difference in FM kg between HED and MED was −2.3 kg [95% CI −5.0 to 0.3]; p=0.09) (Table 2).

#### Fat-free mass

The DCON and MED group decreased in FFM compared to CON. In contrast, HED FFM did not change compared to CON. Due to the unchanged values in FFM kg in HED, the HED group increased compared to DCON and tended to increase compared to MED. There was no difference in FFM kg between MED and DCON (Table 2). There was a trend for a dose-dependent effect for FFM kg (linear trend p=0.004).

All intervention groups increased in FFMpp compared to CON (p<0.001 for comparison for HED and MED, and p=0.001 for DCON) in a dose-dependent manner (linear trend p<0.0001). MED and HED increased FFMpp compared to DCON. There was no difference in FFMpp between HED and MED (Table 2).

#### Visceral adipose tissue volume

All intervention groups reduced VAT cm^3^ compared to CON (DCON vs. CON −666 cm^3^ [95% CI −1109 to −313], MED vs. CON −1264 cm^3^ [95% CI −2129 to −725], and HED vs. CON −1786 cm^3^ [95% CI −2381 to −1132]) in a dose-dependent manner (linear trend [95% CI −800.28 to −480.45]). The reduction from baseline corresponds to −1, −16, −30, and −42% for CON, DCON, MED, and HED, respectively. MED and HED reduced VAT cm^3^ compared to DCON (MED vs. DCON −598 cm^3^ [95% CI −1444 to −9], and HED vs. DCON −1120 cm^3^ [95% CI −1665 to −332]). The difference in VAT cm^3^ between HED and MED was −522 cm^3^ [95% CI −1364 to 403] (Table 2).

### Post hoc outcomes

No changes were observed in leg FFM kg in the intervention groups compared to CON (Table 2). However, MED and HED increased leg FFM kg compared to DCON with no difference between MED and HED. All intervention groups decreased aSAT cm^3^ compared to CON in a dose-dependent manner (linear trend 95% CI: −811 to −266), however, no difference between intervention groups (Table 2). All intervention groups increased in FFM:FM ratio compared to CON in a dose-dependent manner (linear trend p<0.0001). MED and HED increased in FFM:FM ratio compared to DCON and there was no difference between HED and MED (Table 2). There was no indication of a difference in VAT/aSAT in any group (Table 2).

### Sensitivity analyses

A per-protocol sensitivity analysis generally confirmed the intention-to-treat findings (Supplementary Table S10).

### Mediation analyses

Insulin sensitivity and beta-cell DI were mediated by the changes in FM%, FM kg, and body weight in all intervention groups. No body composition variables mediated insulin secretion rate (Supplementary Table S11).

## Discussion

All interventions decreased FMpp more than standard care and adding exercise three or six times/per week to a caloric restriction decreased FM% more than caloric restriction alone in a dose-dependent manner. Furthermore, adding exercise to caloric restriction seemed to prevent the loss in FFM observed with caloric restriction alone.

The literature is mixed on the effects of adding exercise to a hypocaloric diet on weight loss. Our results are in line with previous findings that suggests adding exercise to caloric restriction generally augments weight loss but with diminishing return of benefit with increasing exercise doses on weight loss [30]. However, our results contrast with studies showing no significant additional effect on weight loss or fat loss when exercise is added to a hypocaloric diet [19]. For example, another 16-week study observed no effect on weight loss when adding aerobic training five times/per week or resistance training three times/per week (total 3 sets/per week) to a ∼1000 kcal/day caloric restriction on weight loss compared to caloric restriction alone (10-13% weight loss). Although, FFM preservation and a small, non-significant, fat loss was observed in the exercising groups [31]. Likewise, a 12-week very-low-calorie-diet (VLCD) combined with aerobic training three times/per week demonstrated overall similar reduction compared to the VLCD alone in weight loss (∼12.5 kg/11%), FM, and VAT (∼30%) [18]. A third study comparing 500-1000 kcal/day caloric restriction to caloric restriction plus aerobic training three times/per week also observed no differences in VAT, aSAT, or total abdominal adipose tissue concurrent to a 5% weight loss [17]. These suggests that exercise may not be effective at providing additional weight loss when combined with small or large caloric deficits. Or that the exercise doses/modalities were not sufficient to create a significant net negative energy balance and/or to preserve FFM so that the weight loss was primarily from fat.

The VAT reduction relative to weight loss in the MED - and DCON groups are generally akin to other studies adding exercise to caloric restriction or implementing caloric restriction alone [4, 18, 31]. However, HED seems to lead to a larger reduction in VAT relative to weight loss; this may be explained by the greater FM reduction in HED. This is in line with the proposed allometric relationship of VAT/FM which is consistent regardless of energy restriction modality, and essentially means that it is the fat loss that dictates VAT loss and not weight loss *per sé* [32]. This underlines the importance of aiming at losing FM when undertaking a hypocaloric diet.

The contrasting findings in FFM and FM between our and other studies may be partly explained by the exercise modalities and doses. In support of this, a 6-month intervention of aerobic training 4-5 days/per week and resistance training (4-6 sets/per week) combined with 500-1000 kcal/day caloric restriction greatly reduced body weight and FM while preserving FFM compared to caloric restriction only [33]. Moreover, 6-months of 500-750 kcal/day caloric restriction demonstrated notable differences in FM, VAT, and FFM with equivalent weight loss (∼10%) combined with either three times/per week of aerobic training, resistance training (3-9 sets/per week), or a combination. The overall largest improvements were caloric restriction plus resistance- and aerobic training combined [16]. Impressively, resistance training three times/per week (9-15 sets/per week) completely preserved FFM following a 12-week VLCD (800 kcal/day) supplemented with 80 grams protein/day (total 1120 kcal/day), while the VLCD only group lost 4.6 kg FFM. Both VLCD plus resistance training and VLCD only lost ∼15 kg of FM but the proportion of FM was significantly larger in VLCD plus resistance training (96% loss from FM and 4% from FFM) compared to VLCD only (76% loss from FM and 24% from FFM) [12]. These results suggest that implementing sufficient resistance training, and protein intake, is effective at preserving FFM under large caloric deficits, and provide further fat loss when combined with aerobic training during smaller caloric deficits.

In the present study MED increased more in FFMpp compared to DCON but both MED and DCON lost equal amount of FFM kg, whereas HED completely preserved FFM kg. The differences in FFM kg between MED and HED may partly relate to the resistance training dose. The MED group performed resistance training/upper-body training at 3 sets once/per week, while HED performed 3 sets twice/per week. This is in line with that resistance training ≤10 sets/per week exerts a dose-dependent increase in skeletal muscle hypertrophy somewhat independent of weekly training frequency [34]. Resistance training ∼4 sets/per week seems sufficient for skeletal muscle hypertrophy [35] and while more sets/per week generally promotes more skeletal muscle hypertrophy, 12-20 sets/per week is likely most effective, but these numbers of sets/per week were not examined when undertaking a catabolic state like weight loss [36]. Aerobic training (especially high intensity) may itself promote a degree of FFM preservation [37], which may explain why there was no difference in leg FFM kg between MED and HED. Together, these results suggests that resistance training >3 sets/per week for the major muscle groups are on average required to fully preserved FFM during a 25% caloric deficit. Noteworthy, our participants’ starting point of a relatively high FM, high daily protein intake in the intervention groups, and low physical activity levels/novelty to training in the intervention groups, may increase the chances of preserving FFM during caloric deficits [14, 15]. Practically, resistance training may be regarded as FFM preserving whereas aerobic training and physical activity facilitates a net negative energy balance [15, 16, 30, 37, 38]. Implementing sufficient sets/per week of resistance training to preserve FFM while also performing aerobic training appears most effective for maximizing fat loss during smaller caloric deficits. During larger caloric deficits (e.g., ∼1000 kcal daily deficits, VLCD) resistance training may be superior to aerobic training in maximizing the proportion of fat loss in total weight loss. Importantly, the beneficial physiological adaptations of resistance - and aerobic training exist on a spectrum and may easily be combined [37, 39]. In the present study, it is plausible that going from exercising three to six times/per week was sufficient to completely preserve FFM, and together with a higher exercise energy expenditure, resulted in greater FM and VAT reductions relative to weight loss in HED compared to MED.

### Limitations

There are limitations to our study. *First*. We assessed dietary adherence via a self-reported 3-day diary which may entail information bias, but self-reported dietary adherence was overall similar between intervention groups. *Second*. As this is a secondary analysis, we may not have sufficient statistical power and may increase the risk of type 2 errors. *Third*. Although intervention-related adverse events were low, it cannot be excluded that exercising six times/per week was too much for some individuals to both recover from and adapt to, which may have dampened exercise performance and FFM preservation and added more variation to an inherent individual response to exercise [40].

## Conclusion

Findings from this study suggest that exercise six times/per week result in greater FM reduction than exercise three times/per week, possibly through higher exercise energy expenditure. Moreover, doubling the volume of exercise seems to completely prevent the loss of FFM otherwise observed with diet-induced weight loss.

## Acknowledgments

We would like to thank the study participants for their time and engagement in this study and current and former staff at the Centre for Physical Activity Research, Rigshospitalet. Also, the current and former staff at the Centre for Diabetes Research at the Municipality of Copenhagen for their recruitment support. Lastly, we thank Karen Kettless and Else Danielsen for their technical support and guidance.

## Funding/Support

The project was supported by a grant from TrygFonden and Svend Andersen Fonden. The Centre for Physical Activity Research is supported by TrygFonden (grants ID 101390, ID 20045, and ID 125132). Mark Preben Printz Lyngbaek was supported by a research grant from the Danish Diabetes Academy, which is funded by the Novo Nordisk Foundation, grant number NNF17SA0031406. Cody Durrer was supported by the Canadian Institutes of Health Research (MFE-176582).

## Conflict of Interest

TPA owns stocks in NovoNordisk A/S. TPJS owns an academic consulting business (Blazon Scientific) and an endurance and nutrition consulting business (Veohtu) — these companies had no control over the research design, data analysis, or publication outcomes of this work.

## Author Contributions and Guarantor Statement

MPPL and MRL conceived the study. MPPL wrote the first draft. MPPL, GEL, MRL, TPA, KK, BKP, TPJS, MAVL, CD, BL, CE, and CL contributed to protocol development and study design. NSN, BL GEL, and MPPL performed the experiments and collected the data. CD and NSN performed the DXA and MRI analysis. CE screened the MRI scans for incidental pathology. MRL, CD, NSN, and MPPL integrated and quality-checked the data. CD performed the statistical analyses. MPPL, CD, and MRL wrote the statistical analysis plan. All authors critically read and revised for important intellectual content and approved the final version of the manuscript. MRL is the Principal Investigator. MRL and MPPL are the guarantors and had full access to the data in the study, verified the data, and take responsibility for the integrity of the data and the accuracy of the analysis. MRL and MPPL had full responsibility for the decision to submit and publish.

## Notes

### Clinical Trial

NCT03769883

### Author Declarations

The study, which this study is a secondary analysis of, was approved by the Scientific Ethical Committee of the Capital Region of Denmark (approval number H-18038298).

