## Supplemental Material for "Adding Different Exercise Training Volumes to a Diet-Induced Weight Loss Facilitates Fat Loss and Maintains Fat-Free Mass in a Dose-Depending Fashion in Persons with Newly Diagnosed Type 2 Diabetes: Secondary Findings from the DOSE-EX Multi-Arm, Parallel-Group, Randomized Trial"

**Online-Only Supplemental Material**

**Supplementary Table 1.**

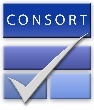
CONSORT 2010 checklist of information to include when reporting a randomised trial*

| Section/Topic | Item No | Checklist item | Reported on page No |
| --- | --- | --- | --- |
| Title and abstract | | | |
|  | 1a | Identification as a randomised trial in the title | 1 |
|  | 1b | Structured summary of trial design, methods, results, and conclusions (for specific guidance see CONSORT for abstracts) | 2 |
| Introduction | | | |
| Background and objectives | 2a | Scientific background and explanation of rationale | 4-5 |
|  | 2b | Specific objectives or hypotheses | 4-5 |
| Methods | | | |
| Trial design | 3a | Description of trial design (such as parallel, factorial) including allocation ratio | 5-10 |
|  | 3b | Important changes to methods after trial commencement (such as eligibility criteria), with reasons | Statistical analyses plan |
| Participants | 4a | Eligibility criteria for participants | 5 |
|  | 4b | Settings and locations where the data were collected | 5 |
| Interventions | 5 | The interventions for each group with sufficient details to allow replication, including how and when they were actually administered | 5-6 |
| Outcomes | 6a | Completely defined pre-specified primary and secondary outcome measures, including how and when they were assessed | 5-8 |
|  | 6b | Any changes to trial outcomes after the trial commenced, with reasons | N/A |
| Sample size | 7a | How sample size was determined | 8 |
|  | 7b | When applicable, explanation of any interim analyses and stopping guidelines | N/A |
| Randomisation: |  |  | 8 |
| Sequence generation | 8a | Method used to generate the random allocation sequence |  |
|  | 8b | Type of randomisation; details of any restriction (such as blocking and block size) | 8, protocol paper |
| Allocation concealment mechanism | 9 | Mechanism used to implement the random allocation sequence (such as sequentially numbered containers), describing any steps taken to conceal the sequence until interventions were assigned | 8, protocol paper |
| Implementation | 10 | Who generated the random allocation sequence, who enrolled participants, and who assigned participants to interventions | 8, protocol paper |
| Blinding | 11a | If done, who was blinded after assignment to interventions (for example, participants, care providers, those assessing outcomes) and how | 8, protocol paper |
|  | 11b | If relevant, description of the similarity of interventions | N/A |
| Statistical methods | 12a | Statistical methods used to compare groups for primary and secondary outcomes | 8-10 |
|  | 12b | Methods for additional analyses, such as subgroup analyses and adjusted analyses | 8-10 |
| Participant flow (a diagram is strongly recommended) | 13a | For each group, the numbers of participants who were randomly assigned, received intended treatment, and were analysed for the primary outcome | 10-11 |
|  | 13b | For each group, losses and exclusions after randomisation, together with reasons | 10-11 |
| Recruitment | 14a | Dates defining the periods of recruitment and follow-up | 5 |
|  | 14b | Why the trial ended or was stopped | N/A |
| Baseline data | 15 | A table showing baseline demographic and clinical characteristics for each group | 22-23 |
| Numbers analysed | 16 | For each group, number of participants (denominator) included in each analysis and whether the analysis was by original assigned groups | 22-23 |
| Outcomes and estimation | 17a | For each primary and secondary outcome, results for each group, and the estimated effect size and its precision (such as 95% confidence interval) | 25 |
|  | 17b | For binary outcomes, presentation of both absolute and relative effect sizes is recommended | N/A |
| Ancillary analyses | 18 | Results of any other analyses performed, including subgroup analyses and adjusted analyses, distinguishing pre-specified from exploratory | Supplementary Table 2.  Supplementary table 10 |
| Harms | 19 | All important harms or unintended effects in each group (for specific guidance see CONSORT for harms) | Supplementary table 13 |
| Limitations | 20 | Trial limitations, addressing sources of potential bias, imprecision, and, if relevant, multiplicity of analyses | 17 |
| Generalisability | 21 | Generalisability (external validity, applicability) of the trial findings | 17 |
| Interpretation | 22 | Interpretation consistent with results, balancing benefits and harms, and considering other relevant evidence | 17 |
| Other information | | | 5 |
| Registration | 23 | Registration number and name of trial registry |  |
| Protocol | 24 | Where the full trial protocol can be accessed, if available | With submission |
| Funding | 25 | Sources of funding and other support (such as supply of drugs), role of funders | 18 |

*We strongly recommend reading this statement in conjunction with the CONSORT 2010 Explanation and Elaboration for important clarifications on all the items. If relevant, we also recommend reading CONSORT extensions for cluster randomised trials, non-inferiority and equivalence trials, non-pharmacological treatments, herbal interventions, and pragmatic trials. Additional extensions are forthcoming: for those and for up to date references relevant to this checklist, see [www.consort-statement.org](http://www.consort-statement.org)

**Supplementary Table 2. Adherence to diet intervention.**

|  |  | **Baseline** |  | **Week 4** | **% adherence** |  | **Week 12** | **% adherence** |  | **Week 16** | **% adherence** | **% adherence after randomization** | **Mean reduction after randomization (Kcal/day)** | **Within +/- 30% of diet plan (N(%))** |
| --- | --- | --- | --- | --- | --- | --- | --- | --- | --- | --- | --- | --- | --- | --- |
|  | n | N= 81 (98.8) | n | N= 79 (96.3) |  | n | N= 75 (91.5) |  | n | N= 74 (90.2) |  |  |  |  |
| **Total energy intake (Kcal/day)** |  |  |  |  |  |  |  |  |  |  |  |  |  |  |
| CON | 20 (100.0) | 2181.3 (711.9) | 17 (85.0) | 1927.0 (1773.0 to 2225.0) | N/A | 17 (85.0) | 1696.0 (1363.0 to 1974.0) | N/A | 17 (85.0) | 1696.0 (1508.0 to 2349.0) | N/A | N/A | -207.4 (495.3) | N/A |
| DCON | 21 (100.0) | 2151.6 (530.2) | 21 (100.0) | 1320.0 (1296.0 to 1486.0) | 90.0 (10.4) | 21 (100.0) | 1535.0 (1344.0 to 1666.0) | 96.0 (13.3) | 21 (100.0) | 1415.0 (1152.0 to 1613.0) | 90.6 (19.9) | 92.2 (10.7) | -707.9 (452.1) | 21 (100.0) |
| MED | 19 (95.0) | 2039.4 (459.3) | 20 (100.0) | 1449.0 (1337.0 to 1653.0) | 90.8 (18.4) | 18 (90.0) | 1499.5 (1382.0 to 1747.0) | 95.6 (23.7) | 17 (85.0) | 1493.0 (1372.0 to 1776.0) | 91.5 (18.2) | 90.8 (18.4) | -509.0 (424.2) | 18 (90.0) |
| HED | 21 (100.0) | 2150.6 (638.7) | 21 (100.0) | 1485.0 (1373.0 to 1804.0) | 87.6 (13.1) | 19 (90.5) | 1460.0 (1357.0 to 1794.0) | 87.0 (13.2) | 19 (90.5) | 1494.0 (1294.0 to 1772.0) | 89.3 (14.6) | 87.6 (13.1) | -619.5 (527.5) | 19 (90.5) |
| **Total carbohydrate (% of total energy intake)** |  |  |  |  |  |  |  |  |  |  |  |  |  |  |
| CON | 20 (100.0) | 41.6 (6.8) | 17 (85.0) | 44.5 (8.6) | 7 (41.2) | 17 (85.0) | 42.5 (5.7) | 9 (52.9) | 17 (85.0) | 40.4 (36.3 to 44.4) | 4 (23.5) | 6.7 (33.3) | NA |  |
| DCON | 21 (100.0) | 42.6 (8.9) | 21 (100.0) | 42.1 (7.4) | 10 (47.6) | 21 (100.0) | 40.9 (7.9) | 6 (28.6) | 21 (100.0) | 42.1 (35.9 to 46.5) | 6 (28.6) | 7.3 (34.9) | NA |  |
| MED | 19 (95.0) | 39.0 (9.0) | 20 (100.0) | 43.5 (4.8) | 8 (40.0) | 18 (90.0) | 45.3 (5.0) | 8 (44.4) | 17 (85.0) | 46.1 (37.7 to 49.0) | 9 (52.9) | 8.3 (41.7) | NA |  |
| HED | 21 (100.0) | 42.3 (8.5) | 21 (100.0) | 42.9 (5.9) | 9 (42.9) | 19 (90.5) | 43.1 (7.1) | 7 (36.8) | 19 (90.5) | 42.8 (39.4 to 45.3) | 5 (26.3) | 7.0 (33.3) | NA |  |
| **Fiber (% of total energy intake)** |  |  |  |  |  |  |  |  |  |  |  |  |  |  |
| CON | 20 (100.0) | 2.0 (1.6 to 2.8) | 17 (85.0) | 2.1 (1.6 to 2.6) | NA | 17 (85.0) | 1.8 (1.6 to 2.5) | NA | 17 (85.0) | 2.0 (1.8 to 2.4) | NA | NA | NA |  |
| DCON | 21 (100.0) | 2.1 (1.7 to 2.5) | 21 (100.0) | 3.8 (3.5 to 4.4) | NA | 21 (100.0) | 3.8 (3.1 to 4.0) | NA | 21 (100.0) | 3.7 (2.9 to 4.4) | NA | NA | NA |  |
| MED | 19 (95.0) | 2.1 (1.6 to 3.2) | 20 (100.0) | 3.9 (3.3 to 4.9) | NA | 18 (90.0) | 3.9 (3.6 to 4.3) | NA | 17 (85.0) | 3.8 (3.3 to 4.4) | NA | NA | NA |  |
| HED | 21 (100.0) | 2.1 (1.7 to 2.6) | 21 (100.0) | 3.7 (3.2 to 4.4) | NA | 19 (90.5) | 3.4 (3.0 to 4.1) | NA | 19 (90.5) | 3.2 (2.8 to 4.0) | NA | NA | NA |  |
| **Total fat (% of total energy intake)** |  |  |  |  |  |  |  |  |  |  |  |  |  |  |
| CON | 20 (100.0) | 37.1 (5.7) | 17 (85.0) | 38.0 (29.6 to 41.8) | 7 (41.2) | 17 (85.0) | 37.0 (34.4 to 40.7) | 5 (29.4) | 17 (85.0) | 37.4 (32.1 to 42.7) | 7 (41.2) | 6.3 (31.7) | NA |  |
| DCON | 21 (100.0) | 36.2 (8.8) | 21 (100.0) | 32.4 (27.6 to 34.7) | 15 (71.4) | 21 (100.0) | 34.9 (30.9 to 40.7) | 11 (52.4) | 21 (100.0) | 32.8 (27.5 to 40.6) | 10 (47.6) | 12.0 (57.1) | NA |  |
| MED | 19 (95.0) | 38.3 (10.7) | 20 (100.0) | 30.3 (27.0 to 36.3) | 14 (70.0) | 18 (90.0) | 32.3 (27.7 to 35.0) | 14 (77.8) | 17 (85.0) | 32.4 (23.9 to 36.6) | 11 (64.7) | 13.0 (65.0) | NA |  |
| HED | 21 (100.0) | 35.2 (7.1) | 21 (100.0) | 32.2 (26.8 to 35.4) | 13 (61.9) | 19 (90.5) | 31.4 (27.7 to 35.4) | 14 (73.7) | 19 (90.5) | 35.7 (30.0 to 39.0) | 8 (42.1) | 11.7 (55.6) | NA |  |
| **Saturated fat (% of total energy intake)** |  |  |  |  |  |  |  |  |  |  |  |  |  |  |
| CON | 20 (100.0) | 12.0 (3.1) | 17 (85.0) | 13.6 (11.7 to 14.7) | 1 (5.9) | 17 (85.0) | 13.5 (12.1 to 15.2) | 1 (5.9) | 17 (85.0) | 12.6 (11.1 to 13.8) | 1 (5.9) | 1.0 (5.0) | NA |  |
| DCON | 21 (100.0) | 12.4 (3.3) | 21 (100.0) | 7.4 (6.0 to 9.9) | 9 (42.9) | 21 (100.0) | 8.9 (7.3 to 10.3) | 5 (23.8) | 21 (100.0) | 8.5 (7.2 to 10.5) | 5 (23.8) | 6.3 (30.2) | NA |  |
| MED | 19 (95.0) | 14.0 (5.4) | 20 (100.0) | 8.9 (7.0 to 11.0) | 6 (30.0) | 18 (90.0) | 8.0 (6.5 to 8.8) | 5 (27.8) | 17 (85.0) | 7.5 (6.6 to 11.5) | 6 (35.3) | 5.7 (28.3) | NA |  |
| HED | 21 (100.0) | 12.1 (3.5) | 21 (100.0) | 7.9 (7.3 to 9.7) | 4 (19.0) | 19 (90.5) | 7.9 (6.8 to 11.1) | 5 (26.3) | 19 (90.5) | 9.4 (7.8 to 12.4) | 3 (15.8) | 4.0 (19.0) | NA |  |
| **Protein (% of total energy intake)** |  |  |  |  |  |  |  |  |  |  |  |  |  |  |
| CON | 20 (100.0) | 18.0 (2.8) | 17 (85.0) | 15.3 (14.4 to 18.6) | 6 (35.3) | 17 (85.0) | 18.0 (15.4 to 18.7) | 11 (64.7) | 17 (85.0) | 19.2 (16.0 to 21.9) | 8 (47.1) | 8.3 (41.7) | NA |  |
| DCON | 21 (100.0) | 18.2 (2.8) | 21 (100.0) | 22.7 (20.8 to 24.2) | 3 (14.3) | 21 (100.0) | 20.5 (17.5 to 23.3) | 8 (38.1) | 21 (100.0) | 21.3 (20.3 to 23.6) | 4 (19.0) | 5.0 (23.8) | NA |  |
| MED | 19 (95.0) | 19.1 (2.3) | 20 (100.0) | 21.4 (19.8 to 23.9) | 5 (25.0) | 18 (90.0) | 21.3 (19.4 to 22.5) | 5 (27.8) | 17 (85.0) | 21.5 (19.6 to 24.7) | 5 (29.4) | 5.0 (25.0) | NA |  |
| HED | 21 (100.0) | 18.2 (3.3) | 21 (100.0) | 22.1 (20.6 to 22.9) | 4 (19.0) | 19 (90.5) | 22.1 (18.7 to 24.8) | 8 (42.1) | 19 (90.5) | 21.8 (18.5 to 23.9) | 7 (36.8) | 6.3 (30.1) | NA |  |
| **Alcohol (% of total energy intake)** |  |  |  |  |  |  |  |  |  |  |  |  |  |  |
| CON | 20 (100.0) | 0.0 (0.0 to 2.6) | 17 (85.0) | 0.0 (0.0 to 0.0) | NA | 17 (85.0) | 0.0 (0.0 to 2.8) | NA | 17 (85.0) | 0.0 (0.0 to 0.0) | NA | NA | NA |  |
| DCON | 21 (100.0) | 0.0 (0.0 to 1.3) | 21 (100.0) | 0.0 (0.0 to 0.0) | NA | 21 (100.0) | 0.0 (0.0 to 0.0) | NA | 21 (100.0) | 0.0 (0.0 to 0.0) | NA | NA | NA |  |
| MED | 19 (95.0) | 0.0 (0.0 to 1.3) | 20 (100.0) | 0.0 (0.0 to 0.0) | NA | 18 (90.0) | 0.0 (0.0 to 0.0) | NA | 17 (85.0) | 0.0 (0.0 to 0.0) | NA | NA | NA |  |
| HED | 21 (100.0) | 0.0 (0.0 to 2.3) | 21 (100.0) | 0.0 (0.0 to 0.0) | NA | 19 (90.5) | 0.0 (0.0 to 0.0) | NA | 19 (90.5) | 0.0 (0.0 to 0.0) | NA | NA | NA |  |

**Supplementary Table 2. Adherence to diet intervention.** CON: Control group, DCON: Diet control group, MED: Moderate exercise dose, HED: High exercise dose. Data is mean (standard deviation) or median (interquartile range).

**Supplementary Table 3. Dose of aerobic training.**

|  | **Minutes pr. week** | **Minutes completed from prescribed (%)** | **Minutes completed within target HR (%)** | **Minutes pr. session** | **Sessions pr. week** |
| --- | --- | --- | --- | --- | --- |
| **Week 1-2** Familiarization |  |  |  |  |  |
| MED | 143 (14) | 95 (10) | 97 (13) | 51 (3) | 3 (0) |
| HED | 208 (48) | 93 (12) | 99 (4) | 45 (4) | 5 (1) |
| **Week 3-10** |  |  |  |  |  |
| MED | 144 (23) | 96 (15) | 99 (2) | 50 (1) | 3 (1) |
| HED | 268 (79) | 89 (27) | 100 (0) | 50 (3) | 5 (2) |
| **Week 11-16** |  |  |  |  |  |
| MED | 138 (34) | 92 (22) | 100 (2) | 49 (1) | 3 (0) |
| HED | 257 (91) | 86 (30) | 99 (3) | 50 (1) | 6 (1) |
| **Week 3-16** |  |  |  |  |  |
| MED | 142 (27) | 94 (18) | 99 (1) | 50 (1) | 3 (1) |
| HED | 264 (84) | 88 (28) | 100 (1) | 50 (3) | 5 (2) |

**Supplementary Table 3. Dose of aerobic training.** Data are mean (SD), HRmax: Maximum heart rate, MED: Moderate exercise dose, HED: High exercise dose.

**Supplementary Table 4. Intensity (internal and external load) in aerobic training.**

|  | **Average relative heart rate (%HRmax)** | **Minutes within 60-79% HRmax** | **Minutes within 80-100% HRmax** | **Relative duration within 80-100% HRmax (%)** | **Average watt** |
| --- | --- | --- | --- | --- | --- |
| **Week 1-2** Familiarization |  |  |  |  |  |
| MED | 73 (4) | 231 (55) | 49 (50,5) | 17 (18) | 69 (27) |
| HED | 71 (3) | 364 (96) | 46 (27,8) | 11 (6) | 73 (24) |
| **Week 3-10** |  |  |  |  |  |
| MED | 76 (2) | 806 (173) | 327 (136) | 29 (11) | 93 (32) |
| HED | 75 (2) | 1554 (491) | 572 (315) | 26 (12) | 111 (46) |
| **Week 11-16** |  |  |  |  |  |
| MED | 79 (4) | 502 (129) | 364 (137) | 42 (16) | 117 (39) |
| HED | 78 (3) | 977 (285) | 714 (248) | 43 (14) | 150 (60) |
| **Week 3-16** |  |  |  |  |  |
| MED | 77 (3) | 1282 (325) | 672 (282) | 34 (12) | 103 (35) |
| HED | 76 (2) | 2438 (866) | 1218 (617) | 32 (13) | 124 (52) |

**Supplementary Table 4. Intensity (internal and external load) in aerobic training.** Data are mean (SD), HRmax: Maximum heart rate, MED: Moderate exercise dose, HED: High exercise dose.

**Supplementary Table 5. Dose of resistance training in the large muscle groups.**

|  | **Sets pr. week Mean (SD)** | **Sets completed from prescribed (%)** | **Sets completed within target RIR (%)** |
| --- | --- | --- | --- |
| **Week 1-2** Familiarization week |  |  |  |
| **MED** |  |  |  |
| Leg press | 2.8 (0.7) | 92.5 (24.5) | 91.2 (11.6) |
| Leg extension | 2.8 (0.8) | 92.9 (26.7) | 100 (0) |
| Leg curl | 2.5 (1.2) | 83.3 (40.8) | 90 (14.9) |
| Chest press | 2.7 (0.9) | 90 (30.8) | 99.1 (3.9) |
| Back row | 2.7 (0.9) | 90 (30.8) | 92.6 (8.5) |
| Total | 10.9 (3.5) | 90.6 (28.9) | 95.2 (5.1) |
| **HED** |  |  |  |
| Leg press | 5.7 (0.8) | 95.2 (12.8) | 90.9 (8.3) |
| Leg extension | 5.9 (0.4) | 97.9 (7.2) | 95.4 (6.8) |
| Leg curl | 5.3 (1.1) | 88.9 (18.2) | 92 (7.7) |
| Chest press | 5.7 (0.8) | 95.2 (12.8) | 96.7 (5.1) |
| Back row | 5.6 (0.8) | 94 (13.5) | 96.6 (10.2) |
| Total | 22.7 (3) | 94.6 (12.7) | 94.5 (5.2) |
| **Week 3-10** |  |  |  |
| **MED** |  |  |  |
| Leg press | 2.9 (0.5) | 96.9 (17.1) | 89.9 (3.8) |
| Leg extension | 3 (0.1) | 99.1 (3.3) | 94 (3.9) |
| Leg curl | 2.7 (1) | 89.6 (32) | 87.9 (4.6) |
| Chest press | 2.9 (0.5) | 96.9 (17.1) | 90.6 (4.3) |
| Back row | 2.9 (0.5) | 96.9 (17.1) | 90.3 (4.4) |
| Total | 11.6 (2.1) | 96.6 (17.2) | 91.5 (3.2) |
| **HED** |  |  |  |
| Leg press | 5.1 (1.7) | 85.1 (28.4) | 91.5 (5.2) |
| Leg extension | 5.9 (0.2) | 97.9 (4.1) | 94.5 (2.3) |
| Leg curl | 4 (2.2) | 67.4 (37.3) | 90 (5.3) |
| Chest press | 5.1 (1.7) | 85.4 (28.6) | 93.9 (4.7) |
| Back row | 5.1 (1.7) | 85.4 (28.6) | 93.2 (4.8) |
| Total | 20.4 (6.8) | 85.2 (28.5) | 92.8 (4) |
| **Week 11-16** |  |  |  |
| **MED** |  |  |  |
| Leg press | 2.9 (0.7) | 95.8 (22.9) | 94.2 (6) |
| Leg extension | 3 (0) | 100 (0) | 93.3 (5.4) |
| Leg curl | 2.6 (1.3) | 86.1 (42.7) | 94.8 (5.2) |
| Chest press | 2.9 (0.7) | 95.8 (22.9) | 94.5 (5.2) |
| Back row | 2.9 (0.7) | 95.8 (22.9) | 93.9 (5.1) |
| Total | 11.5 (2.7) | 94.7 (23.2) | 94.1 (3.9) |
| **HED** |  |  |  |
| Leg press | 5.3 (1.9) | 87.8 (31.8) | 94.2 (3.4) |
| Leg extension | 6 (0.1) | 100.7 (2.4) | 95.4 (2.1) |
| Leg curl | 4.3 (2.7) | 72.2 (44.9) | 96 (3.2) |
| Chest press | 5.3 (1.9) | 88.5 (31.9) | 97 (2.6) |
| Back row | 5.3 (1.9) | 88.5 (31.9) | 96.1 (2.7) |
| Total | 21.2 (7.6) | 88.3 (31.8) | 95.7 (1.5) |
| **Week 3-16** |  |  |  |
| **MED** |  |  |  |
| Leg press | 2.9 (0.6) | 96.4 (19.5) | 91.5 (3.7) |
| Leg extension | 3 (0.1) | 99.5 (1.9) | 93.7 (3.1) |
| Leg curl | 2.6 (1.1) | 88.1 (36.3) | 89.9 (4.1) |
| Chest press | 2.9 (0.6) | 96.1 (19.5) | 93.7 (3.6) |
| Back row | 2.9 (0.6) | 96.4 (19.5) | 91.6 (3.1) |
| Total | 11.6 (2.3) | 96.2 (19.4) | 92.3 (2.7) |
| **HED** |  |  |  |
| Leg press | 5.2 (1.8) | 86.3 (29.5) | 92.5 (3.7) |
| Leg extension | 5.9 (0.1) | 99.1 (1.6) | 94.9 (1.7) |
| Leg curl | 4.2 (2.4) | 69.4 (40.2) | 92 (4.8) |
| Chest press | 5.2 (1.8) | 86.7 (29.6) | 94.9 (3.8) |
| Back row | 5.2 (1.8) | 86.7 (29.6) | 94.5 (2.6) |
| Total | 20.8 (7.1) | 86.5 (29.6) | 93.9 (2.6) |

**Supplementary Table 5. Dose of resistance training in the large muscle groups**. Data are mean (SD), RIR: repetitions in reserve, MED: Moderate exercise dose, HED: High exercise dose.

**Supplementary Table 6. External load in resistance training in the large muscle groups.**

|  | **Repetitions pr. week** | **Repetitions pr. set** | **Average kilogram lifted pr. set** | **Tonnage pr. week** |
| --- | --- | --- | --- | --- |
| **Week 1-2** familiarization |  |  |  |  |
| **MED** |  |  |  |  |
| Leg press | 32.9 (9.8) | 12 (0) | 97.9 (26.3) | 3525 (945.1) |
| Leg extension | 33.4 (9.6) | 12 (0) | 31.5 (9) | 1135.4 (323) |
| Leg curl | 30 (14.7) | 12 (0) | 24.8 (4.9) | 891 (175.4) |
| Chest press | 32.4 (11.1) | 12 (0) | 19.7 (8) | 710 (289.4) |
| Back row | 32.4 (11.1) | 12 (0) | 34.6 (10.1) | 1244.5 (363.1) |
| Total | 130.1 (43) | 12 (0) | 45.5 (11.2) | 6547 (1616.4) |
| **HED** |  |  |  |  |
| Leg press | 69.9 (20.7) | 11.9 (0.5) | 104.7 (22.6) | 7269.9 (2375.8) |
| Leg extension | 69.1 (9.1) | 12 (0.1) | 35.7 (10.3) | 2445 (745.2) |
| Leg curl | 69 (32.4) | 12 (0) | 26.4 (9.7) | 1831 (1059.7) |
| Chest press | 68.7 (21.9) | 11.7 (0.7) | 23.7 (9.9) | 1587.2 (733.3) |
| Back row | 69.6 (20.8) | 12 (0.2) | 41.8 (11.1) | 2869.9 (1051.5) |
| Total | 277.2 (83.6) | 11.9 (0.2) | 50.5 (12.2) | 13908.9 (4727.5) |
| **Week 3-10** |  |  |  |  |
| **MED** |  |  |  |  |
| Leg press | 30 (10.1) | 11.1 (0.4) | 117.6 (33.8) | 3834.4 (1405.2) |
| Leg extension | 32.7 (2.2) | 11.1 (0.3) | 35.8 (11.4) | 1166.1 (386.3) |
| Leg curl | 29.3 (13.7) | 11.1 (0.4) | 58.2 (65.1) | 1891.4 (2202.9) |
| Chest press | 31.7 (7.4) | 11.1 (0.3) | 24 (13.8) | 790.8 (495.4) |
| Back row | 32.1 (7.3) | 11.1 (0.3) | 44.4 (10.7) | 1446.3 (516.2) |
| Total | 125.5 (29.1) | 11.1 (0.3) | 57.2 (17) | 7466.7 (2781.9) |
| **HED** |  |  |  |  |
| Leg press | 52 (20.7) | 11.1 (0.3) | 141.7 (40.7) | 8095.2 (3807) |
| Leg extension | 63.1 (4.8) | 11 (0.2) | 42.2 (11.8) | 2668.6 (778.9) |
| Leg curl | 38 (25) | 11.2 (0.5) | 33.6 (10.1) | 1560.8 (992.6) |
| Chest press | 53.1 (20.9) | 11 (0.3) | 27.3 (11.4) | 1592 (834.8) |
| Back row | 53.5 (21.1) | 11.1 (0.3) | 53.3 (15) | 3109.5 (1312.5) |
| Total | 210.9 (82.7) | 11.1 (0.3) | 65.3 (18.4) | 15022.2 (6628.9) |
| **Week 11-16** |  |  |  |  |
| **MED** |  |  |  |  |
| Leg press | 25.9 (6.7) | 8.9 (0.8) | 140.6 (32.4) | 3768.3 (985.3) |
| Leg extension | 26.9 (2.7) | 9 (0.9) | 44.1 (14.3) | 1148.8 (359.7) |
| Leg curl | 23.5 (12) | 8.8 (0.2) | 65.3 (49.4) | 1799.1 (1247.6) |
| Chest press | 25.8 (6.8) | 8.9 (0.8) | 25.6 (8) | 681.1 (215.3) |
| Back row | 25.9 (6.7) | 8.9 (0.8) | 53.5 (11.5) | 1431 (329.3) |
| Total | 103.3 (27) | 8.9 (0.8) | 67.4 (14.4) | 7209.9 (1669.6) |
| **HED** |  |  |  |  |
| Leg press | 43 (17.1) | 8.7 (0.1) | 153 (59.5) | 7432.3 (3512.4) |
| Leg extension | 50.6 (2.8) | 8.7 (0.1) | 50.3 (13.9) | 2540.8 (716.2) |
| Leg curl | 33.9 (23.1) | 8.6 (0.2) | 42 (12.2) | 1887.8 (981.5) |
| Chest press | 43.6 (17.1) | 8.7 (0.1) | 34.4 (14.1) | 1673 (785.4) |
| Back row | 41.8 (16.5) | 8.7 (0.2) | 61.9 (23.4) | 2931.9 (1357.9) |
| Total | 171.8 (67.2) | 8.7 (0.1) | 72.7 (26) | 13946.1 (6290.5) |
| **Week 3-16** |  |  |  |  |
| **MED** |  |  |  |  |
| Leg press | 29.3 (7) | 10.2 (0.6) | 125.5 (34.4) | 3764.2 (1322.3) |
| Leg extension | 30.2 (2) | 10.2 (0.5) | 39.3 (12.4) | 1171.8 (375.7) |
| Leg curl | 26.7 (12.9) | 10.4 (0.8) | 57.7 (58) | 1717.7 (1795.2) |
| Chest press | 29.1 (7) | 10.2 (0.6) | 24.3 (10.6) | 729.6 (355.2) |
| Back row | 29.4 (7) | 10.2 (0.6) | 47.7 (11) | 1418.1 (478) |
| Total | 117 (27.8) | 10.2 (0.6) | 60.6 (16.5) | 7256 (2512.9) |
| **HED** |  |  |  |  |
| Leg press | 48.7 (18.8) | 10.1 (0.5) | 146.9 (42.7) | 7822 (3533.6) |
| Leg extension | 57.8 (2.3) | 10 (0.1) | 45.7 (12.6) | 2643.9 (743.7) |
| Leg curl | 36.2 (23.9) | 10.3 (0.7) | 36.3 (11.3) | 1617.5 (1053.4) |
| Chest press | 49 (18.9) | 10.1 (0.5) | 29.9 (12.7) | 1612 (849.2) |
| Back row | 49.2 (19) | 10.1 (0.5) | 56.3 (18.3) | 3034.6 (1380.8) |
| Total | 195.4 (75.3) | 10.1 (0.5) | 68.8 (20.2) | 14701.9 (6506.9) |

**Supplementary Table 6. External load in resistance training in the large muscle groups**. Data are mean (SD), Tonnage: weight (kg) x repetitions x sets, MED: Moderate exercise dose, HED: High exercise dose.

**Supplementary Table 7. Exercise modification in aerobic and resistance training.**

|  | **Total number of exercises with modifications stratified by reasons** | | | | | **Relative number with ≥1 modification (%)** | |
| --- | --- | --- | --- | --- | --- | --- | --- |
|  | *Fatigue* | *Musculo-skeletal discomfort* | *Motivational* | *COVID19* | *Other reasons* | *Participants^*^* | *Sessions* |
| **Week 1-16** |  |  |  |  |  |  |  |
| **MED** |  |  |  |  |  |  |  |
| Aerobic training | 1 | 2 | 1 | 0 | 0 | 15 | <1 |
| Leg press | 3 | 2 | 0 | 19 | 6 | 25 | 9 |
| Leg extension | 0 | 0 | 0 | 19 | 0 | 15 | 9 |
| Leg curl | 3 | 2 | 0 | 0 | 0 | 10 | 5 |
| Chest press | 0 | 4 | 0 | 19 | 0 | 30 | 6 |
| Back row | 0 | 0 | 0 | 19 | 0 | 15 | 6 |
| Total resistance training | 7 | 10 | 1 | 76 | 6 | 35 | 7 |
| **HED** |  |  |  |  |  |  |  |
| Aerobic training | 2 | 3 | 1 | 0 | 1 | 24 | <1 |
| Leg press | 0 | 14 | 6 | 6 | 14 | 29 | 6 |
| Leg extension | 0 | 2 | 0 | 0 | 2 | 10 | 1 |
| Leg curl | 0 | 2 | 0 | 0 | 1 | 10 | <1 |
| Chest press | 0 | 0 | 0 | 0 | 3 | 10 | <1 |
| Back row | 0 | 1 | 0 | 0 | 2 | 10 | <1 |
| Total resistance training | 2 | 22 | 7 | 6 | 23 | 29 | 2 |

**Supplementary Table 7. Exercise modification in aerobic and resistance training.** MED: Moderate exercise dose, HED: High exercise dose. The modifications included either 1) dose reductions in minutes/sets/reps/weight, 2) suboptimal installment of the machines (e.g. lower range of motion in leg press) or 3) exercise substitution targeting the same specific muscle group (e.g. leg press could be substituted with squat). Other reasons: Time constraints, equipment unavailability, illness or not reported. Missing sessions are not included. *: Some patients required modifications for several exercises, therefore the total number of exercises is greater that the number of patients.

**Supplementary Table 8. Adherence for aerobic and resistance training.**

|  | **Aerobic training completed from prescribed (%)** | **Resistance training completed from prescribed (%)** | **Total training completed from prescribed (%)** |  | **Aerobic – and resistance training ≥ 70% of prescribed training (N(%))** | |
| --- | --- | --- | --- | --- | --- | --- |
| **Week 1-2** familiarization |  |  |  |  | **MED** | 19 (95) |
| MED | 93 (17) | 86 (28) | 90 (20) |  | **HED** | 18 (86) |
| HED | 92 (16) | 89 (14) | 90 (13) |  |  |  |
| **Week 3-10** |  |  |  |  |  |  |
| MED | 96 (14) | 91 (16) | 94 (15) |  |  |  |
| HED | 90 (27) | 82 (27) | 86 (27) |  |  |  |
| **Week 11-16** |  |  |  |  |  |  |
| MED | 95 (22) | 93 (22) | 93 (22) |  |  |  |
| HED | 86 (31) | 86 (31) | 86 (31) |  |  |  |
| **Week 3-16** |  |  |  |  |  |  |
| MED | 95 (18) | 92 (19) | 93 (18) |  |  |  |
| HED | 88 (28) | 84 (29) | 86 (28) |  |  |  |

**Supplementary Table 8. Adherence for aerobic and resistance training.** Data are mean (SD), MED: Moderate exercise dose, HED: High exercise dose.

**Supplementary Table 9. The number of DXA and MRI scans and deviations from analysis.**

|  | **CON n (%)** | **DCON n (%)** | **MED n (%)** | **HED n (%)** | **All n (%)** |
| --- | --- | --- | --- | --- | --- |
| **Participants missing DXA scan at baseline** | 0 | 0 | 0 | 0 | 0 |
| **Participants missing DXA scan at follow-up** | 2 (10) | 0 | 1 (5) | 2 (10) | 5 (6) |
| Drop out | 2 (10) | 0 | 1 (5) | 2 (10) | 5 (6) |
| **Participants missing MRI scan at baseline** | 4 (20) | 2 (10) | 2 (10) | 2 (10) | 11 (13) |
| Unsatisfied with group allocation | 1 (5) |  |  |  | 1 (1) |
| Claustrophobia | 1 (5) |  | 1 (5) |  | 2 (2) |
| MRI scanner malfunction | 1 (5) |  |  |  | 1 (1) |
| Metal bullet fragments in the wrist | 1 (5) |  |  |  | 1 (1) |
| Could not fit in MRI scanner |  | 1 (5) | 1 (5) |  | 2 (2) |
| Metal implant in the spine from previous surgery |  | 1 (5) |  |  | 1 (1) |
| **Participants missing MRI scan at follow-up** | 8 (40) | 4 (19) | 3 (15) | 2 (10) | 17 (21) |
| Unsatisfied with group allocation at baseline and therefore no follow-up scan | 1 (5) |  |  |  | 1 (1) |
| Claustrophobia at pre-scan and therefore no follow-up -scan | 1 (5) |  | 1(5) |  | 2 (2) |
| Claustrophobia |  | 1 (5) |  |  | 1 (1) |
| MRI scanner malfunction at baseline-scan and therefore no follow-up -scan | 1 (5) |  |  |  | 1 (1) |
| Metal bullet fragments in the wrist at baseline scan and therefore no follow-up -scan | 1 (5) |  |  |  | 1 (1) |
| Could not fit in MRI scanner at baseline-scan and therefore no follow-up -scan |  | 1 (5) | 1 (5) |  | 2 (2) |
| Metal implant in the spine at baseline scan and therefore no follow-up -scan |  | 1 (5) |  |  | 1 (1) |
| Drop out | 2 (10) |  | 1 (5) | 2 (9.5) | 5 (6) |
| COVID19 lockdown of Radiology department | 1 (5) | 1 (5) |  |  | 2 (2) |
| Unknown | 1 (5) |  |  |  | 1 (1) |
| **Number of SAT measurements at baseline** | 9 (45) | 8 (38) | 8 (40) | 8 (38) | 33 (40) |
| **Number of SAT measurements at follow-up** | 8 (40) | 8 (38) | 8 (40) | 8 (38) | 32 (39) |
| **Number of VAT measurements at baseline** | 15 (75) | 18 (86) | 17 (85) | 17 (81) | 67 (82) |
| **Number of VAT measurements at follow-up** | 11 (55) | 16 (76) | 15 (75) | 16 (76) | 24 (29) |

**Supplementary Table 9. The number of DXA and MRI scans and deviations from analysis.** DXA: Dual-Energy X-ray Absorptiometry, MRI: Magnetic resonance imaging, VAT: Visceral adipose tissue, SAT: Abdominal subcutaneous adipose tissue, CON: Control group n=20, DCON: Diet control group n=21, MED: Moderate exercise dose n=20, HED: High exercise dose n=21.

**Supplementary Table 10. Per-protocol sensitivity analysis on pairwise comparisons in the primary outcome and secondary outcomes.**

|  | **HED vs. CON** |  | **MED vs. CON** |  | **DCON vs. CON** |  | **HED vs. DCON** |  | **MED vs. DCON** | | **HED vs. MED** |  | **Global P** |
| --- | --- | --- | --- | --- | --- | --- | --- | --- | --- | --- | --- | --- | --- |
|  | MD (95% CI) | P | MD (95% CI) | P | MD (95% CI) | P | MD (95% CI) | P | MD (95% CI) | P | MD (95% CI) | P |  |
| **Primary outcome** | | | | | | | | | | | | | |
| Fat mass (pp) | -7.7 (-9.9 to -5.5) | <0.001 | -6.1 (-8.3 to -4.0) | <0.001 | -3.2 (-5.4 to -1.1) | 0.003 | -4.4 (-6.6 to -2.3) | <0.001 | -2.9 (-5.0 to -0.8) | 0.007 | -1.6 (-3.7 to 0.6) | 0.16 | <0.001 |
| **Secondary outcomes** | | | | | | | | | | | | | |
| Body weight (kg) | -11.4 (-14.7 to -8.1) | <0.001 | -10.1 (-13.4 to -6.9) | <0.001 | -6.9 (-10.1 to -3.7) | <0.001 | -4.5 (-7.7 to -1.3) | 0.006 | -3.2 (-6.3 to -0.1) | 0.04 | -1.3 (-4.5 to 2.0) | 0.43 | <0.001 |
| Body weight (% difference) | -11.6 (-14.9 to -8.2) | <0.001 | -10.3 (-13.6 to -7.0) | <0.001 | -7.0 (-10.2 to -3.8) | <0.001 | -4.9 (-8.4 to -1.5) | 0.006 | -3.5 (-6.9 to -0.1) | 0.04 | -1.5 (-5.1 to 2.2) | 0.43 | <0.001 |
| Fat mass (kg) | -11.1 (-13.9 to -8.4) | <0.001 | -8.9 (-11.6 to -6.1) | <0.001 | -5.4 (-8.0 to -2.7) | <0.001 | -5.8 (-8.4 to -3.1) | <0.001 | -3.5 (-6.1 to -0.9) | 0.010 | -2.3 (-5.0 to 0.5) | 0.10 | <0.001 |
| Fat-free mass (kg) (% difference) | -0.1 (-1.9 to 1.8)† | 0.96 | -1.9 (-3.7 to -0.1)† | 0.04 | -2.7 (-4.4 to -0.9)† | 0.003 | 2.7 (0.9 to 4.6)† | 0.004 | 0.8 (-0.9 to 2.6)† | 0.35 | 1.9 (0.0 to 3.7)† | 0.05 | 0.003 |
| Fat-free mass (pp) | 8.0 (5.7 to 10.3) | <0.001 | 6.5 (4.2 to 8.7) | <0.001 | 3.5 (1.3 to 5.7) | 0.002 | 4.6 (2.4 to 6.8) | <0.001 | 3.0 (0.9 to 5.2) | 0.007 | 1.6 (-0.7 to 3.8) | 0.17 | <0.001 |
| VAT (cm^3^) | -1732.6 (-2191.0 to -981.4)‡ | NA | -1234.7 (-2134.7 to -686.1)‡ | NA | -590.3 (-1060.1 to -220.7)‡ | NA | -1142.4 (-1624.9 to -326.5)‡ | NA | -644.4 (-1480.6 to 5.5)‡ | NA | -497.9 (-1260.9 to 464.8)‡ | NA | NA |
| **Post hoc outcomes** | | | | | | | | | | | | | |
| aSAT (cm^3^) | -1736.4 (-3806.8 to -691.2)‡ | NA | -1205.2 (-2399.9 to -567.2)‡ | NA | -936.3 (-1664.9 to -410.3)‡ | NA | -800.1 (-3103.2 to 74.1)‡ | NA | -268.9 (-1304.1 to 302.8)‡ | NA | -531.2 (-2949.9 to 385.1)‡ | NA | NA |
| Leg fat-free mass (kg) (% difference) | 1.4 (-2.0 to 4.9) | 0.43 | 1.5 (-1.8 to 5.0) | 0.37 | -2.3 (-5.5 to 1.0) | 0.16 | 3.8 (0.4 to 7.2) | 0.03 | 3.9 (0.6 to 7.4) | 0.02 | -0.2 (-3.5 to 3.3) | 0.92 | 0.06 |
| Leg fat-free mass (pp) | 0.3 (-0.6 to 1.2) | 0.46 | 0.7 (-0.2 to 1.6) | 0.12 | -0.0 (-0.9 to 0.9) | 0.96 | 0.4 (-0.5 to 1.2) | 0.42 | 0.7 (-0.1 to 1.6) | 0.10 | -0.4 (-1.3 to 0.5) | 0.42 | 0.30 |
| FFM/FM | 43.1 (27.6 to 60.5)† | <0.001 | 34.0 (19.7 to 50.0)† | <0.001 | 15.7 (3.6 to 29.2)† | 0.01 | 23.7 (10.8 to 38.1)† | <0.001 | 15.8 (3.9 to 29.1)† | 0.009 | 6.8 (-4.6 to 19.6)† | 0.25 | <0.001 |
| VAT/SAT (% difference) | -4.0 (-28.5 to 16.5)‡ | NA | -24.7 (-68.6 to 20.6)‡ | NA | 3.5 (-10.0 to 20.6)‡ | NA | -7.2 (-26.5 to 16.4)‡ | NA | -27.2 (-84.2 to 7.3)‡ | NA | 27.5 (-21.9 to 103.3)‡ | NA | NA |

**Supplementary Table 10. Per-protocol sensitivity analysis on pairwise comparisons in the primary outcome and secondary outcomes.** Data are adjusted mean differences and 95% confidence intervals derived from constrained baseline longitudinal analysis via linear mixed models. †Mean difference expressed as a percent difference (ratio of geometric means) from the log-transformed analysis. ‡Bias-corrected and accelerated confidence intervals derived from non-parametric bootstrap analysis (in this case a p-value can’t be obtained). MD: Mean difference, CI: confidence intervals, FFM: fat-free mass, FM: fat mass, CON: control group, DCON: Diet control group, MED: Moderate exercise dose, HED: High exercise dose.

**Supplementary Table 11. Post hoc mediation analysis of the treatment effect of the change in body composition on beta-cell indices.**

|  | **ACME (95% CI)** | **ADE (95% CI)** | **Total effect (95% CI)** | **Proportion mediated (95% CI)** |
| --- | --- | --- | --- | --- |
| **Fat mass percentage** | | | | |
| DCON |  |  |  |  |
| Late-phase disposition index | 0.7 (0.2 to 1.5) | 0.4 (-0.4 to 1.1) | 1.1 (0.6 to 1.8) | 0.6 (0.2 to 1.8) |
| Late-phase insulin sensitivity index | 1.2 (0.3 to 3.6) | -0.1 (-2.3 to 1.2) | 1.2 (0.1 to 2.4) | 1.0 (0.2 to 27.4) |
| Late-phase insulin secretion rate | 0.0 (-0.0 to 0.0) | 0.1 (0.1 to 0.3) | 0.1 (0.1 to 0.2) | 0.0 (-0.3 to 0.3) |
| Oral disposition index | 78.0 (41.6 to 163.1) | 70.5 (-12.5 to 158.1) | 148.5 (91.5 to 229.1) | 0.5 (0.2 to 1.3) |
| Oral insulin sensitivity index | 3.1 (1.4 to 6.2) | -0.4 (-3.5 to 1.4) | 2.7 (1.8 to 3.9) | 1.1 (0.5 to 2.8) |
| MED |  |  |  |  |
| Late-phase disposition index | 1.3 (0.4 to 3.0) | 0.5 (-0.6 to 1.6) | 1.8 (1.0 to 3.1) | 0.7 (0.2 to 1.4) |
| Late-phase insulin sensitivity index | 2.4 (0.6 to 7.3) | 0.3 (-1.9 to 2.1) | 2.7 (1.3 to 6.8) | 0.9 (0.2 to 3.0) |
| Late-phase insulin secretion rate | 0.0 (-0.1 to 0.1) | 0.1 (0.0 to 0.3) | 0.1 (0.1 to 0.2) | 0.1 (-0.5 to 0.8) |
| Oral disposition index | 149.7 (77.3 to 315.7) | 46.7 (-131.4 to 135.6) | 196.3 (119.4 to 268.3) | 0.8 (0.4 to 2.0) |
| Oral insulin sensitivity index | 5.9 (2.4 to 12.9) | -0.4 (-4.1 to 1.8) | 5.5 (3.2 to 11.1) | 1.1 (0.6 to 1.9) |
| HED |  |  |  |  |
| Late-phase disposition index | 1.7 (0.6 to 3.2) | 0.1 (-1.5 to 1.2) | 1.8 (1.1 to 2.7) | 0.9 (0.3 to 2.3) |
| Late-phase insulin sensitivity index | 3.0 (1.0 to 7.8) | -0.5 (-5.3 to 1.7) | 2.5 (1.2 to 4.0) | 1.2 (0.3 to 8.1) |
| Late-phase insulin secretion rate | 0.0 (-0.1 to 0.1) | 0.1 (0.0 to 0.2) | 0.1 (0.1 to 0.2) | 0.1 (-1.0 to 0.8) |
| Oral disposition index | 193.1 (103.3 to 431.2) | 55.9 (-51.7 to 140.6) | 249.0 (172.1 to 414.8) | 0.8 (0.4 to 1.3) |
| Oral insulin sensitivity index | 7.6 (3.4 to 14.4) | -2.4 (-8.7 to 0.9) | 5.2 (3.3 to 8.8) | 1.5 (0.8 to 3.8) |
| **Fat-free mass percentage** | | | | |
| DCON |  |  |  |  |
| Late-phase disposition index | 0.7 (0.2 to 1.6) | 0.4 (-0.4 to 1.1) | 1.1 (0.6 to 1.8) | 0.6 (0.2 to 1.8) |
| Late-phase insulin sensitivity index | 1.3 (0.3 to 3.7) | -0.1 (-2.5 to 1.1) | 1.2 (0.1 to 2.4) | 1.1 (0.2 to 19.5) |
| Late-phase insulin secretion rate | 0.0 (-0.0 to 0.1) | 0.1 (0.1 to 0.2) | 0.1 (0.1 to 0.2) | 0.0 (-0.3 to 0.4) |
| Oral disposition index | 83.1 (45.8 to 173.6) | 65.2 (-22.1 to 150.6) | 148.4 (91.1 to 225.4) | 0.6 (0.3 to 1.4) |
| Oral insulin sensitivity index | 3.2 (1.5 to 6.4) | -0.5 (-3.8 to 1.2) | 2.7 (1.7 to 3.8) | 1.2 (0.5 to 3.1) |
| MED |  |  |  |  |
| Late-phase disposition index | 1.3 (0.4 to 3.2) | 0.5 (-0.7 to 1.5) | 1.8 (1.0 to 3.0) | 0.7 (0.2 to 1.6) |
| Late-phase insulin sensitivity index | 2.4 (0.6 to 7.5) | 0.3 (-2.3 to 2.0) | 2.7 (1.3 to 6.3) | 0.9 (0.2 to 2.5) |
| Late-phase insulin secretion rate | 0.0 (-0.1 to 0.1) | 0.1 (0.0 to 0.3) | 0.1 (0.1 to 0.2) | 0.1 (-0.5 to 0.8) |
| Oral disposition index | 156.3 (80.4 to 325.4) | 39.8 (-129.1 to 133.7) | 196.2 (120.2 to 269.8) | 0.8 (0.4 to 1.9) |
| Oral insulin sensitivity index | 6.0 (2.5 to 13.1) | -0.5 (-4.2 to 1.7) | 5.5 (3.2 to 11.5) | 1.1 (0.6 to 2.1) |
| HED |  |  |  |  |
| Late-phase disposition index | 1.7 (0.6 to 3.3) | 0.1 (-1.5 to 1.2) | 1.8 (1.2 to 2.7) | 0.9 (0.3 to 2.5) |
| Late-phase insulin sensitivity index | 3.0 (0.9 to 7.7) | -0.5 (-5.0 to 1.7) | 2.5 (1.2 to 4.0) | 1.2 (0.3 to 5.8) |
| Late-phase insulin secretion rate | 0.0 (-0.1 to 0.1) | 0.1 (0.0 to 0.2) | 0.1 (0.0 to 0.2) | 0.1 (-1.1 to 0.9) |
| Oral disposition index | 199.2 (107.1 to 417.7) | 49.6 (-58.5 to 133.4) | 248.7 (169.7 to 397.1) | 0.8 (0.5 to 1.3) |
| Oral insulin sensitivity index | 7.7 (3.5 to 14.1) | -2.5 (-8.6 to 0.8) | 5.2 (3.3 to 8.5) | 1.5 (0.8 to 3.8) |
| **Visceral adipose tissue (cm^3^)** | | | | |
| DCON |  |  |  |  |
| Late-phase disposition index | 0.3 (-0.3 to 1.0) | 0.5 (-0.5 to 1.5) | 0.8 (0.3 to 1.6) | 0.4 (-0.4 to 2.2) |
| Late-phase insulin sensitivity index | 0.5 (-0.4 to 2.3) | 0.1 (-1.7 to 1.7) | 0.7 (-0.3 to 1.9) | 0.8 (-2.0 to 29.1) |
| Late-phase insulin secretion rate | 0.0 (-0.0 to 0.1) | 0.2 (0.0 to 0.3) | 0.2 (0.1 to 0.3) | 0.1 (-0.3 to 0.6) |
| Oral disposition index | 103.1 (56.3 to 203.8) | 32.4 (-74.0 to 112.5) | 135.5 (71.8 to 225.8) | 0.8 (0.3 to 2.2) |
| Oral insulin sensitivity index | 2.6 (0.7 to 5.3) | -0.4 (-3.4 to 1.7) | 2.2 (1.2 to 3.4) | 1.2 (0.3 to 3.9) |
| MED |  |  |  |  |
| Late-phase disposition index | 0.6 (-0.5 to 1.9) | 1.3 (0.1 to 3.0) | 1.9 (0.9 to 3.4) | 0.3 (-0.4 to 1.0) |
| Late-phase insulin sensitivity index | 1.0 (-0.7 to 4.3) | 1.9 (0.0 to 5.7) | 2.9 (1.2 to 8.3) | 0.3 (-0.5 to 1.2) |
| Late-phase insulin secretion rate | 0.0 (-0.1 to 0.2) | 0.1 (-0.0 to 0.3) | 0.2 (0.1 to 0.3) | 0.2 (-0.4 to 1.1) |
| Oral disposition index | 187.4 (97.1 to 411.8) | 2.8 (-216.6 to 104.4) | 190.2 (101.4 to 280.6) | 1.0 (0.5 to 2.9) |
| Oral insulin sensitivity index | 4.7 (1.1 to 9.7) | 1.3 (-2.6 to 5.0) | 6.0 (3.1 to 14.1) | 0.8 (0.2 to 1.9) |
| HED |  |  |  |  |
| Late-phase disposition index | 0.8 (-0.8 to 2.3) | 1.3 (-0.2 to 2.7) | 2.0 (1.2 to 3.1) | 0.4 (-0.5 to 1.2) |
| Late-phase insulin sensitivity index | 1.3 (-0.9 to 5.5) | 1.5 (-2.5 to 4.0) | 2.8 (1.3 to 4.3) | 0.4 (-0.4 to 2.2) |
| Late-phase insulin secretion rate | 0.0 (-0.1 to 0.2) | 0.1 (-0.1 to 0.2) | 0.1 (0.0 to 0.2) | 0.3 (-1.0 to 1.8) |
| Oral disposition index | 239.1 (121.4 to 533.8) | 38.6 (-120.2 to 146.5) | 277.7 (180.5 to 449.8) | 0.9 (0.5 to 1.5) |
| Oral insulin sensitivity index | 6.0 (1.5 to 12.7) | -0.0 (-6.2 to 3.7) | 6.0 (3.7 to 10.1) | 1.0 (0.3 to 2.4) |
| **Abdominal subcutaneous adipose tissue (cm^3^)** | | | | |
| DCON |  |  |  |  |
| Late-phase disposition index | 1.0 (-0.1 to 6.4) | 0.2 (-3.4 to 1.8) | 1.2 (0.3 to 3.2) | 0.8 (-0.5 to 20.6) |
| Late-phase insulin sensitivity index | 2.5 (0.2 to 13.1) | -1.0 (-10.4 to 1.5) | 1.5 (0.1 to 5.2) | 1.7 (-2.3 to 57.5) |
| Late-phase insulin secretion rate | -0.0 (-0.2 to 0.1) | 0.1 (-0.0 to 0.3) | 0.1 (-0.1 to 0.2) | -0.3 (-6.8 to 0.9) |
| Oral disposition index | 59.2 (-23.6 to 294.9) | 38.4 (-132.2 to 189.7) | 97.6 (23.9 to 237.4) | 0.6 (-0.7 to 6.2) |
| Oral insulin sensitivity index | 4.2 (1.2 to 17.2) | -1.7 (-13.3 to 1.4) | 2.5 (1.2 to 5.7) | 1.7 (0.2 to 20.6) |
| MED |  |  |  |  |
| Late-phase disposition index | 1.1 (-0.2 to 7.6) | 1.1 (-1.8 to 3.2) | 2.2 (0.8 to 7.8) | 0.5 (-0.2 to 5.6) |
| Late-phase insulin sensitivity index | 2.8 (0.2 to 16.9) | 1.6 (-3.2 to 8.8) | 4.3 (1.6 to 19.7) | 0.6 (-0.2 to 4.7) |
| Late-phase insulin secretion rate | -0.0 (-0.2 to 0.1) | 0.2 (-0.0 to 0.4) | 0.1 (0.0 to 0.3) | -0.3 (-3.6 to 1.2) |
| Oral disposition index | 65.3 (-29.8 to 364.5) | 140.3 (-94.1 to 340.2) | 205.6 (52.4 to 415.9) | 0.3 (-0.3 to 2.6) |
| Oral insulin sensitivity index | 4.7 (1.0 to 22.1) | 5.2 (0.6 to 30.5) | 9.8 (3.4 to 31.8) | 0.5 (0.0 to 1.9) |
| HED |  |  |  |  |
| Late-phase disposition index | 1.8 (-0.1 to 8.7) | 0.5 (-4.2 to 2.8) | 2.3 (1.5 to 5.9) | 0.8 (-0.3 to 7.0) |
| Late-phase insulin sensitivity index | 4.3 (0.5 to 32.7) | -1.2 (-23.5 to 2.4) | 3.2 (1.6 to 8.5) | 1.4 (0.0 to 32.3) |
| Late-phase insulin secretion rate | -0.1 (-0.2 to 0.1) | 0.2 (-0.0 to 0.3) | 0.1 (0.0 to 0.2) | -0.5 (-5.0 to 1.2) |
| Oral disposition index | 101.9 (-38.5 to 436.4) | 55.4 (-265.0 to 204.8) | 157.3 (76.9 to 277.8) | 0.6 (-0.3 to 5.5) |
| Oral insulin sensitivity index | 7.3 (2.3 to 46.0) | -2.7 (-33.5 to 2.4) | 4.6 (2.8 to 9.4) | 1.6 (0.4 to 21.7) |

**Supplementary Table 11. Post hoc mediation analysis of the treatment effect of the change in body composition on beta-cell indices.** Data are marginal means with 95% confidence intervals (CI). ACME: Average causal mediation effect, ADE; Average direct effect. DCON: Diet control group, MED: Moderate exercise dose, HED: High exercise dose.

**Supplementary Table 12. Free-living physical activity.**

|  | **Baseline** | **Week 4** | **Week 12** | **Week 16** |
| --- | --- | --- | --- | --- |
| N participants | 81 | 74 | 73 | 73 |
| **Valid days (N)** |  |  |  |  |
| CON | 6.0 (1.4) | 6.9 (1.1) | 6.5 (0.7) | 5.8 (1.5) |
| DCON | 6.4 (1.3) | 6.9 (0.9) | 6.5 (0.8) | 5.8 (1.5) |
| MED | 5.8 (1.1) | 7.0 (1.2) | 6.3 (0.9) | 6.2 (1.2) |
| HED | 6.0 (1.6) | 6.4 (0.6) | 6.1 (1.1) | 5.5 (1.5) |
| **Non-wear time (min/day)#** |  |  |  |  |
| CON | 0 (0 ; 5) | 2 (0 ; 20) | 0 (0 ; 7) | 0 (0 ; 0) |
| DCON | 0 (0 ; 0) | 0 (0 ; 16) | 0 (0 ; 10) | 0 (0 ; 0) |
| MED | 0 (0 ; 0) | 0 (0 ; 0) | 0 (0 ; 0) | 0 (0 ; 0) |
| HED | 0 (0 ; 0) | 0 (0 ; 7) | 0 (0 ; 0) | 0 (0 ; 0) |
| **Total physical activity (counts per minute)#** |  |  |  |  |
| CON | 291 (215 ; 376) | 303 (248 ; 356) | 321 (224 ; 369) | 277 (174 ; 332) |
| DCON | 276 (224 ; 366) | 256 (185 ; 355) | 236 (184 ; 373) | 251 (210 ; 388) |
| MED | 276 (168 ; 369) | 259 (159 ; 309) | 228 (160 ; 373) | 270 (193 ; 372) |
| HED | 303 (226 ; 340) | 322 (231 ; 521) | 404 (254 ; 519) | 300 (244 ; 456) |
| **MVPA (min/day)#** |  |  |  |  |
| CON | 17 (11 ; 24) | 21 (12 ; 30) | 21 (6 ; 32) | 15 (5 ; 28) |
| DCON | 15 (6 ; 20) | 13 (7 ; 30) | 14 (7 ; 25) | 14 (8 ; 23) |
| MED | 15 (4 ; 27) | 14 (6 ; 30) | 13 (6 ; 22) | 14 (7 ; 26) |
| HED | 14 (8 ; 25) | 28 (7 ; 53) | 29 (13 ; 54) | 24 (16 ; 46) |
| **Sitting time (min/day)#** |  |  |  |  |
| CON | 529 (484 ; 611) | 562 (488 ; 608) | 527 (445 ; 600) | 529 (507 ; 595) |
| DCON | 592 (515 ; 654) | 588 (501 ; 667) | 610 (504 ; 692) | 563 (492 ; 610) |
| MED | 635 (429 ; 684) | 620 (543 ; 670) | 612 (547 ; 724) | 592 (466 ; 668) |
| HED | 580 (514 ; 671) | 534 (452 ; 628) | 580 (463 ; 650) | 598 (552 ; 693) |
| **Stepping (steps/day)#** |  |  |  |  |
| CON | 5499 (3561 ; 7452) | 5157 (3905 ; 7730) | 5764 (3985 ; 7182) | 4478 (3201 ; 7063) |
| DCON | 4303 (3855 ; 5798) | 4758 (3907 ; 7109) | 4125 (3192 ; 6247) | 4708 (3164 ; 6522) |
| MED | 4034 (2953 ; 6216) | 4195 (3025 ; 5191) | 4268 (2316 ; 5768) | 4180 (3170 ; 5021) |
| HED | 4380 (3336 ; 7110) | 5190 (3510 ; 9686) | 7025 (5204 ; 8562) | 5398 (3967 ; 9514) |

**Supplementary Table 12. Free-living physical activity**. Baseline data are means and standard deviations unless stated otherwise. #Median and interquartile range (p25 to p75). CON: Control group, DCON: Diet control group, MED: Moderate exercise dose, HED: High exercise dose, MVPA: Moderate and vigorous physical activity.

**Supplementary Table 13. Adverse events after randomization.**

|  | **CON n (%)** | **DCON n (%)** | **MED n (%)** | **HED n (%)** | **All n (%)** |
| --- | --- | --- | --- | --- | --- |
| **Serious AE** | 2 (10.0) | 1 (4.8) | 0 (0.0) | 0 (0.0) | 3 (3.7) |
| **Infections (COVID19)** | 0 (0.0) | 0 (0.0) | 2 (10.0) | 0 (0.0) | 2 (2.4) |
| **Musculoskeletal pain and discomfort** |  |  |  |  |  |
| Back pain | 0 (0.0) | 0 (0.0) | 0 (0.0) | 1 (4.8) | 1 (1.2) |
| Lower extremities | 0 (0.0) | 0 (0.0) | 0 (0.0) | 0 (0.0) | 0 (0.0) |
| Upper extremities | 0 (0.0) | 0 (0.0) | 0 (0.0) | 1 (4.8) | 1 (1.2) |
| Other | 0 (0.0) | 0 (0.0) | 0 (0.0) | 2 (9.5) | 2 (2.4) |
| **Musculoskeletal injury** |  |  |  |  |  |
| Back pain | 0 (0.0) | 0 (0.0) | 0 (0.0) | 1 (4.8) | 1 (1.2) |
| Lower extremities | 0 (0.0) | 0 (0.0) | 1 (5.0) | 1 (4.8) | 2 (2.4) |
| Fatigue | 0 (0.0) | 0 (0.0) | 0 (0.0) | 1 (4.8) | 1 (1.2) |
| Other | 0 (0.0) | 0 (0.0) | 0 (0.0) | 1 (4.8) | 1 (1.2) |
| **Complications associated with clinical or experimental procedures** |  |  |  |  |  |
| Allergic reactions to bandages and wound plasters | 0 (0.0) | 0 (0.0) | 1 (5.0) | 2 (9.5) | 3 (3.7) |
| Felt uncomfortable during hyperglycemic clamp | 0 (0.0) | 1 (4.8) | 1 (5.0) | 0 (0.0) | 2 (2.4) |
| Pain from muscle biopsy | 0 (0.0) | 1 (4.8) | 1 (5.0) | 0 (0.0) | 2 (2.4) |
| Peripheral intravenous catheter went subcutaneous during hyperglycemic clamp | 0 (0.0) | 1 (4.8) | 0 (0.0) | 0 (0.0) | 1 (1.2) |
| **Nutrition** | 0 (0.0) | 0 (0.0) | 0 (0.0) | 0 (0.0) | 0 (0.0) |

**Supplementary Table 13. Adverse events after randomization.** Numerator: Number of adverse events in each group, Denominator: Total number of participants in each group. *Numerator is the number of participants with at least one event in each group and denominator is the total number of participants in the group.

SAE: Serious Adverse Event, AE: Adverse Event, CON: Control group n= 20, DCON: Diet control group n= 21, MED: Moderate exercise dose n= 20, HED: High exercise dose n= 21. Serious AE included one case of transient ischemic attack and one malignant melanoma in the CON group and one case of prolactinoma in DCON group. Musculoskeletal pain and discomfort: Defined as musculoskeletal pain or discomfort but the ability to modify an exercise so that the prescribed exercise intervention can be performed. Musculoskeletal injury: An injury is defined as pain or discomfort to an extend that precludes participating in at least one protocol prescribed exercise >7 days. Nutrition: Events of increased or decreased hunger or satiety.

**Supplementary Figure 1: Changes from constrained baseline to 16 weeks follow-up in the primary and secondary outcomes.**

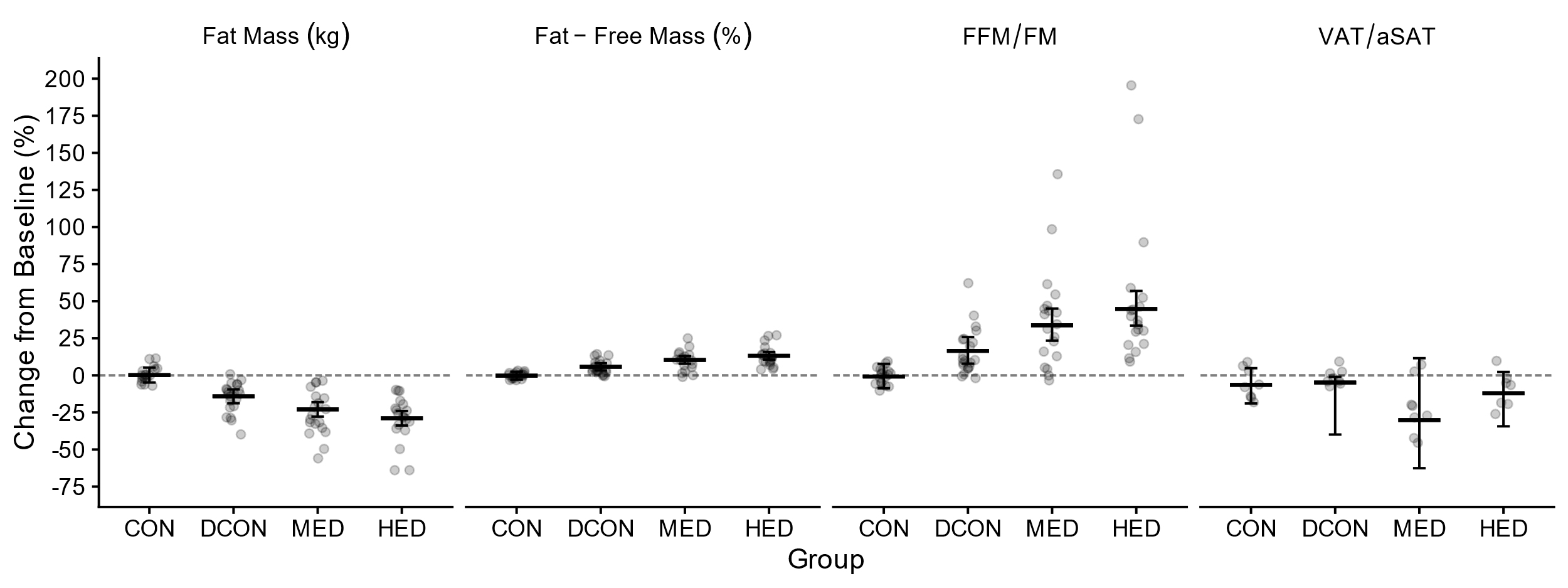
**Supplementary Figure 1: Changes from constrained baseline to 16 weeks follow-up in the primary and secondary outcomes.** Data are presented as marginal means (bar charts overlaid with individual values) with 95% confidence intervals. Data were analyzed using a constrained baseline longitudinal model via linear mixed models. Results are adjusted for sex. †Mean difference expressed as a percent difference (ratio of geometric means) from log-transformed analysis. ‡Bias-corrected and accelerated confidence intervals derived from non-parametric bootstrap analysis (in this case a p-value can’t be obtained). FFM/FM†; fat-free mass to fat mass ratio. VAT/aSAT‡: Visceral adipose tissue to abdominal subcutaneous adipose tissue ratio. Fat mass (kg), Fat-free mass (pp). CON: Control group, DCON: Diet control group, MED: Moderate exercise dose, HED: High exercise dose.
